# Deep Immune Profiling of MIS-C demonstrates marked but transient immune activation compared to adult and pediatric COVID-19

**DOI:** 10.1101/2020.09.25.20201863

**Authors:** Laura A. Vella, Josephine R. Giles, Amy E. Baxter, Derek A. Oldridge, Caroline Diorio, Leticia Kuri-Cervantes, Cécile Alanio, M. Betina Pampena, Jennifer E. Wu, Zeyu Chen, Yinghui Jane Huang, Elizabeth M. Anderson, Sigrid Gouma, Kevin O. McNerney, Julie Chase, Chakkapong Burudpakdee, Jessica H. Lee, Sokratis A. Apostolidis, Alexander C. Huang, Divij Mathew, Oliva Kuthuru, Eileen C. Goodwin, Madison E. Weirick, Marcus J. Bolton, Claudia P. Arevalo, Andre Ramos, Cristina Jasen, Heather M. Giannini, Kurt D’Andrea, The UPenn COVID Processing Unit, Nuala J. Meyer, Edward M. Behrens, Hamid Bassiri, Scott E. Hensley, Sarah E. Henrickson, David T. Teachey, Michael R. Betts, E. John Wherry

## Abstract

Pediatric COVID-19 following SARS-CoV-2 infection is associated with fewer hospitalizations and often milder disease than in adults. A subset of children, however, present with Multisystem Inflammatory Syndrome in Children (MIS-C) that can lead to vascular complications and shock, but rarely death. The immune features of MIS-C compared to pediatric COVID-19 or adult disease remain poorly understood. We analyzed peripheral blood immune responses in hospitalized SARS-CoV-2 infected pediatric patients (pediatric COVID-19) and patients with MIS-C. MIS-C patients had patterns of T cell-biased lymphopenia and T cell activation similar to severely ill adults, and all patients with MIS-C had SARS-CoV-2 spike-specific antibodies at admission. A distinct feature of MIS-C patients was robust activation of vascular patrolling CX3CR1+ CD8 T cells that correlated with use of vasoactive medication. Finally, whereas pediatric COVID-19 patients with acute respiratory distress syndrome (ARDS) had sustained immune activation, MIS-C patients displayed clinical improvement over time, concomitant with decreasing immune activation. Thus, non-MIS-C versus MIS-C SARS-CoV-2 associated illnesses are characterized by divergent immune signatures that are temporally distinct and implicate CD8 T cells in clinical presentation and trajectory of MIS-C.

**One Sentence Summary:** MIS-C is defined by generalized lymphocyte activation that corrects during hospitalization, including elevated plasmablast frequencies and marked activation of vascular patrolling CX3CR1+ CD8 T cells.

## INTRODUCTION

Coronavirus disease 19 (COVID-19) from severe acute respiratory syndrome coronavirus 2 (SARS-CoV-2) infection is primarily an illness of adults, with morbidity and mortality increased with advanced age ^1–6^. In contrast, COVID-19 hospitalization rates are low in children, with fewer than 0.1% of the total deaths occurring in those under 21 years of age ^7–10^. The underlying reasons for differences in pediatric versus adult SARS-CoV-2 infection remain unclear given that other respiratory viruses, including influenza and respiratory syncytial virus, can cause substantial morbidity and mortality in young children as well as in older adults ^11–16^.

In adults, COVID-19 morbidity is associated with increases in clinical markers of inflammation, including C-reactive protein (CRP), lactic dehydrogenase (LDH), ferritin, D-dimer, and procalcitonin (PCT) ^2,5^. Clinical measures of cellular immunity are limited, but decreased absolute lymphocyte counts (ALC) are a consistent finding and are associated with worse outcomes ^17–21^. Translational studies in adult COVID-19 identified T cell lymphopenia in particular, with a preferential decrease in CD8 T cells compared to CD4 T cells ^21–23^. In those lymphocytes remaining, marked activation of CD4 and CD8 T cells was observed, and T cell activation correlated positively with severity of illness ^22,24–28^. However, even among the sickest adults, the degree of immune activation varies ^22,24,25,29^.

Reports of children hospitalized with COVID-19 suggest similar clinical inflammatory profiles, including mild to moderate elevations of CRP, ferritin, PCT, and reduced ALC ^30–32^. However, how these soluble inflammatory markers relate to cellular immune perturbations is not known, as clinical laboratory tests do not include cellular measures of immune activation and dysfunction. It therefore remains unclear whether the outcome differences in children compared to adults are associated with distinct profiles of cellular immune activation, involvement of different immune cell types, or if differences are non-immunological and low pediatric mortality occurs despite a similar immune landscape.

After the initial wave of COVID-19 hospitalizations, children began to develop Multisystem Inflammatory Syndrome in Children (MIS-C), a syndrome that commonly presents with vascular involvement and shock but is rarely fatal ^33–40^. MIS-C has been suggested to be a post-infectious or delayed-infectious event ^37,39^ and has similarities in clinical presentation to Kawasaki disease, especially the vascular involvement. However, MIS-C and Kawasaki disease differ in key clinical, inflammatory, and autoantibody signatures ^37,39,41,42^. Initial studies suggested that MIS-C was associated with similar or higher levels of clinical inflammatory markers as observed in adult and pediatric COVID-19 ^34–36,43^. However, clinically, MIS-C has a distinct presentation compared to the respiratory manifestations of more typical pediatric or adult COVID-19. The immunologic features driving MIS-C presentation remain poorly defined.

To understand immune responses during SARS-CoV-2 illness in children, including acute SARS-CoV-2 infection (pediatric COVID-19) and MIS-C, we collected peripheral blood samples from patients admitted to the Children’s Hospital of Philadelphia in April through June of 2020. We performed high dimensional flow cytometry in parallel with samples collected from adults admitted with COVID-19, as well as healthy adults and those recovered from documented SARS-CoV-2 infection, as recently described ^22,25^. These cellular analyses were paired with serologic and plasma cytokine data and integrated with clinical and laboratory information. Our results demonstrate that although the immune landscape in pediatric COVID-19 is similar to adults, MIS-C represents an exacerbated T cell activation state, particularly for CD8 T cells, including a highly activated vascular patrolling CD8 T cell subset, and follows a distinct temporal immunological trajectory. Specifically, these features of profound T cell activation decrease concurrently with clinical improvement. Together, our findings provide a broad immunologic foundation for understanding pathogenesis and recovery in this novel COVID-19 associated inflammatory syndrome in children with potential implications for adult COVID-19 patients.

## RESULTS

### Cytopenias in MIS-C include enhanced T cell lymphopenia

To interrogate the immunologic features of pediatric SARS-CoV-2 infection, we collected peripheral blood from patients admitted to the Children’s Hospital of Philadelphia from April through July 2020. Patients were initially approached for consent after testing positive for SARS-CoV-2 by polymerase chain reaction (PCR). In May, enrollment criteria were expanded to include any patient diagnosed with MIS-C, regardless of PCR status (see Methods for MIS-C clinical inclusion criteria). Fifty-seven patients were enrolled during the study period, and 30 of the 57 patients had samples collected and processed for cellular immunophenotyping. Of the 30 included patients, 16 were diagnosed with COVID-19 and 14 were diagnosed with MIS-C (**Fig 1A, Supplementary Table 1**). Of the patients with COVID-19, 4 were determined to have acute respiratory distress syndrome (ARDS). The remaining COVID-19 patients were classified as non-ARDS and included those intubated for lung-extrinsic disease or respiratory pathology not consistent with ARDS; those requiring non-invasive ventilation; patients requiring nasal cannula alone; or patients who were identified as SARS-CoV-2 positive incidentally during routine screening procedures. Patients with MIS-C were treated with intravenous immunoglobulin (IVIg) and most were treated with steroids (**Supplementary Table 1**). Consistent with previous reports ^34–36,43^, the maximum values for clinical measures of inflammation (including LDH, ferritin, CRP, etc) varied but were elevated in most subjects (**Fig 1B**). Although inflammatory markers in COVID-19 and MIS-C were comparable, measures of inflammation were more often obtained for patients admitted to the intensive care unit (ICU) and for patients with MIS-C; as a result, patients with less severe COVID-19 and possibly lower inflammation are underrepresented (**Supplementary Table 1**). We next investigated relationships between clinical markers of inflammation, demographics such as age, sex, and race as well as clinical disease measures including the need for vasoactive medications and hospitalization length. As expected, clinical measures of inflammation tended to correlate with one another (**Fig 1C**), but the clinical diagnosis of MIS-C, while less likely in someone with other comorbidities, was not distinguished from pediatric COVID-19 (ARDS or non-ARDS) by correlation with individual laboratory measures, with the exception of hyponatremia, which was nominally correlated with MIS-C (**Fig 1D**). Further, the white blood cell counts with differentials were variable (**Fig S1A, 1B**), as was the lymphocyte nadir (**Fig 1E**). Nevertheless, both the ALC and platelet nadir did have nominal correlations with clinical disease metrics consistent with severe illness (i.e. with hospitalization length and vasoactive requirement, respectively) (**Fig 1D**). A low ALC was associated with higher D-dimer as well as a longer hospitalization length, and a low ALC was less likely in pediatric COVID-19 subjects who did not have ARDS. A low platelet count was nominally correlated with D-dimer and ARDS as well as with PCT, CRP, and requirement of vasoactive medications (e.g. epinephrine). Taken together, these clinical values suggested heterogeneity in pediatric patients with COVID-19 and MIS-C, with inflammatory marker elevation and cytopenias occurring in both groups.

**Figure 1.**
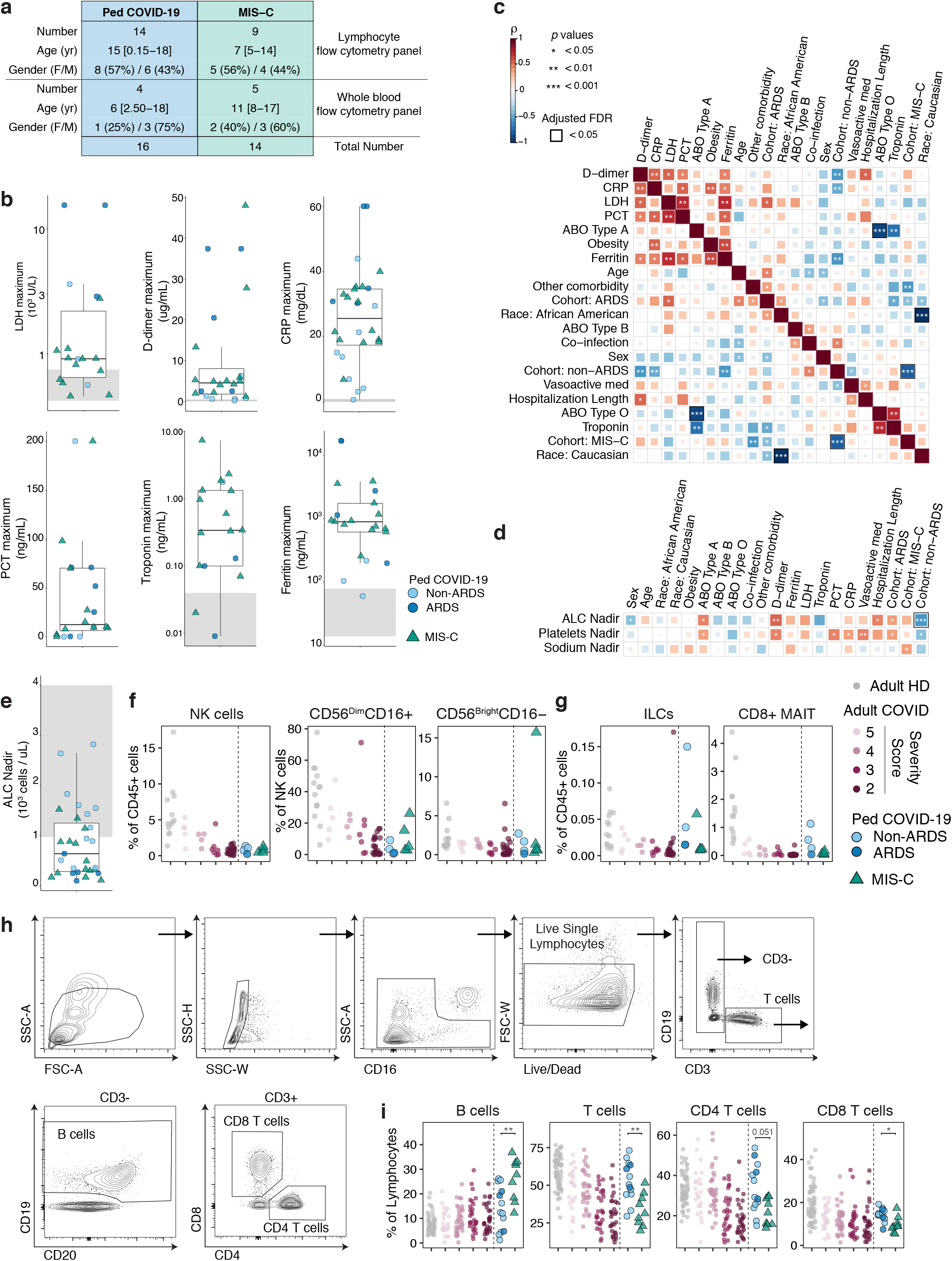
Cytopenias in MIS-C include enhanced T cell lymphopenia. (**A**) Overview of pediatric cohorts in study, including Pediatric COVID-19 (Ped COVID-19) and MIS-C. (**B**) Maximum clinical inflammatory markers in the pediatric cohorts. (**C**) Spearman correlation and hierarchical clustering of indicated clinical features and maximum laboratory values for pediatric cohorts. (**D**) Spearman correlation and hierarchical clustering of clinical parameters with lowest recorded values (nadir) for indicated features for pediatric cohorts. (**E**) Nadir absolute lymphocyte counts (ALC) for pediatric cohorts. (**F**) Frequencies of total NK cells and NK subsets across adult healthy donors (HD), adult COVID-19, and pediatric cohorts. (**G**) Frequencies of ILCs (left) and CD8+ MAIT cells (right) across all cohorts. (**H**) Gating scheme for major lymphocyte populations; B cells, CD4, and CD8 T cells. (**I**) Quantification of major lymphocyte populations across all cohorts. (**BEFGI**) each dot represents an individual patient or subject, with adult HD in grey circles, adult COVID-19 in shades of mauve indicated by disease severity score, Ped COVID-19 in blue circles, with ARDS patients in dark blue, and MIS-C in green triangles. (**BE**) Normal laboratory reference ranges for healthy pediatric subjects are indicated in grey shading. (**CD**) Spearman’s Rank Correlation coefficient (ρ) is indicated by square size and heat scale; significance indicated by: * p<0.05, ** p<0.01, and *** p<0.001; black box indicates FDR<0.05. (**FGI**) significance determined by unpaired Wilcoxon test between Ped COVID-19 and MIS-C groups only, indicated by: * p<0.05, ** p<0.01.

To further understand leukocyte biology in pediatric SARS-CoV-2 infection, we performed high dimensional flow cytometry on whole blood or freshly isolated peripheral blood mononuclear cells (PBMC) (**Fig 1A, Supplementary Tables 2 and 3**) in parallel with adult samples from a recently reported dataset that were re-analyzed for integration with pediatric data^22,25^. As a result, we were able to compare the immunologic phenotype between pediatric MIS-C and pediatric COVID-19 within the context of healthy adults and the spectrum of adult COVID-19 severity.

We first assessed non-lymphocytes in whole blood by identifying monocytes, neutrophils, eosinophils, immature granulocytes, and dendritic cells (DC) (**Fig S1C**). As a proportion of CD45+ cells, eosinophils and immature granulocytes were similar between pediatric COVID-19 and MIS-C. Neutrophils tended to be increased in MIS-C by flow cytometric and clinical differential measurements, though this change was not statistically significant (**Fig S1D, S1B**). Total monocytes and DC as well as subsets of each were also comparable (**Fig S1A, S1E, S1F**). However, plasmacytoid DC were decreased in MIS-C (**Fig S1F**), consistent with whole blood observations in a MIS-C cohort from New York ^44^ and data from adults ^24,25,45^. As these flow cytometric analyses are relative rather than absolute, frequency changes observed may reflect the lymphopenia in 9 of 14 MIS-C patients at the time of research blood draw (**Fig S1B**).

Transient lymphopenia is a well-established feature of viral infection ^46–50^ but is typically only observed around the time of symptom onset and then recovers quickly ^51^. The features that lead to transient lymphopenia in humans during acute viral infections are poorly understood and may depend on the infection or host factors. Reduced ALC has also been noted in adult COVID-19, and more severe lymphopenia is associated with worse clinical outcome ^17–22^. In our cohort, lymphopenia was observed in most patients diagnosed with MIS-C or ARDS (**Fig 1E**), consistent with previous studies ^30,32,34–36,43,52^. We ^22^ and others ^21,23^ have recently noted that in adult COVID-19 patients, lymphopenia is T cell biased, with a preferential decrease in CD8 T cells. However, relative impact on lymphocyte subsets and phenotype in pediatric COVID-19 and MIS-C has not been defined. We, therefore, interrogated B cells, CD4 T cells, CD8 T cells, mucosal associated invariant T cells (MAITs), and natural killer (NK) cells using high dimensional flow cytometry. Total NK cell and NK subset frequencies were reduced in both pediatric cohorts compared to healthy adults, consistent with findings in adult COVID-19 (**Fig 1F, Fig S1C**). Although CD8+ MAITs were decreased in both pediatric cohorts compared to healthy adults, innate lymphocyte (ILC) frequencies appeared similar to healthy adults (**Fig S1C, Fig 1G**) ^25^. Unlike adult COVID-19, pediatric COVID-19 with or without ARDS was not consistently associated with reduced T cell frequencies (**Fig 1H, Fig 1I**, dark blue circles). In contrast, patients with MIS-C displayed reduced T cell frequencies compared to healthy adults, particularly for CD8 T cells. It is possible that these comparisons could differ slightly if compared in parallel to flow cytometry performed on freshly isolated samples from healthy pediatric subjects, but recruitment of healthy pediatric subjects was not permitted during the study period. Nevertheless, these data demonstrate that pediatric COVID-19 and MIS-C are both inflammatory disease presentations with concomitant lymphopenia, though the T cell bias of the lymphopenia in MIS-C is more similar to severely ill adults.

### Increased T cell activation in MIS-C compared to pediatric COVID-19

To investigate T cell responses in our pediatric versus adult cohorts in more detail, we first examined age-associated differences. As expected ^53^, the frequency of naive CD4 and CD8 T cells was higher in the pediatric cohorts (**Fig 2A**) and decreased with age (**Fig 2B**). Because of this difference in the proportion of naive T cells, all comparisons of T cell populations focused on non-naive subsets. For both CD4 and CD8 T cells, the frequency of central memory (CM, CD45RA-CD27+CCR7+) and the effector memory 1 (EM1, CD45RA-CD27+CCR7-) subsets was similar between adult and pediatric cohorts, whereas the effector memory 2 (EM2, CD45RA-CD27-) subset was slightly lower in children (**Fig S2A-C**). The terminally differentiated effector memory subset that re-expresses CD45RA (EMRA, CD45RA+CD27-CCR7-) increases with age ^54–58^. Thus, as expected, EMRA were present at a lower frequency in our pediatric cohorts even when only examining non-naive T cells (**Fig S2B, S2C**). The naive, effector, and central memory subset distributions were not different between the pediatric COVID-19 and MIS-C (**Fig S2A-C**).

**Figure 2.**
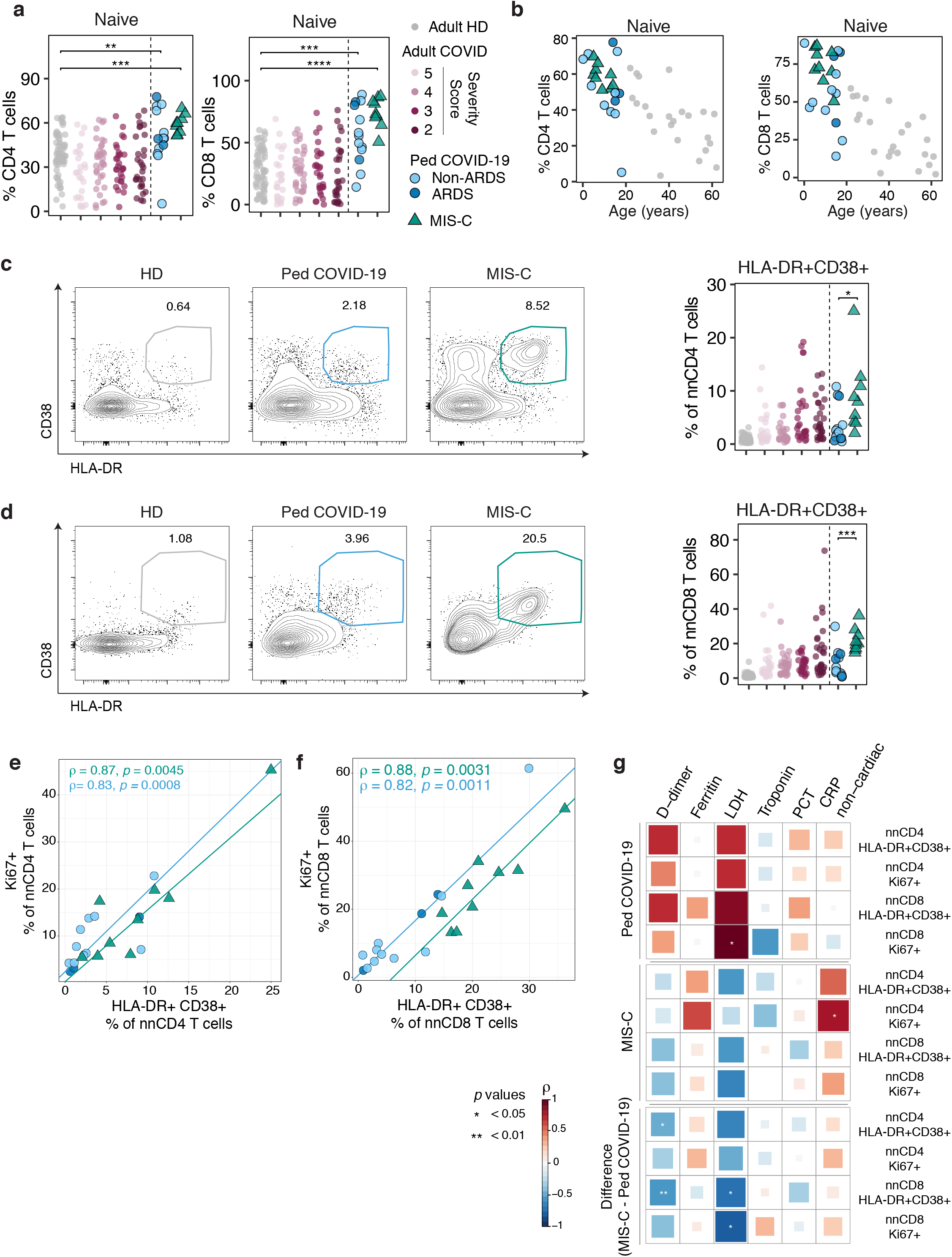
T cell activation in MIS-C is greater than in COVID-19. (**A**) Proportion of naive cells within CD4 (left) and CD8 (right) T cells in all cohorts. For comparison between pediatric cohorts and adult HD, significance was determined by unpaired Wilcoxon test, indicated by: * p<0.05, ** p<0.01, *** p<0.001. (**B**) Association between age (years) and proportion of naive T cells for pediatric cohorts and adult HD. (**C**) Representative flow cytometry plots and quantification of HLA-DR+CD38+ non-naive CD4 T cells (nnCD4 T cells). (**D**) Representative flow cytometry plots and quantification of HLA-DR+CD38+ non-naive CD8 T cells (nnCD8 T cells). (**EF**) Correlation between Ki67+ and HLA-DR+CD38+ populations for nnCD4 (**E**) and nnCD8 (**F**) T cells in pediatric cohorts. Non-parametric trend lines (Sen-Theil) for each pediatric cohort, with Spearman’s Rank Correlation coefficient (ρ) and associated P value shown. (**G**) Spearman correlation for indicated clinical features with frequencies of HLA-DR+CD38+ and Ki67+ nnCD4 and nnCD8 T cells. Top panels indicate correlations within each cohort (Ped COVID-19 and MIS-C). Bottom panel indicates the difference in Spearman correlation between the two pediatric cohorts. Spearman’s Rank Correlation coefficient (ρ) is indicated by square size and heat scale; when calculating the difference in ρ between Ped COVID-19 and MIS-C (**G**, bottom panel), the resulting value was divided by 2 in order to normalize to a −1 to 1 scale; significance indicated by: * p<0.05, ** p<0.01. (**ABCDE**) Each dot represents an individual patient or subject, with adult HD in grey circles, adult COVID-19 in shades of mauve indicated by disease severity score, Ped COVID-19 in blue circles, with ARDS patients in dark blue, and MIS-C in green triangles. (**CD**) Significance determined by unpaired Wilcoxon test between Ped COVID-19 and MIS-C groups only, indicated by: * p<0.05, *** p<0.001.

T cell activation and proliferation are hallmarks of acute viral infections ^59^ and are characteristic features of adult COVID-19 patients ^22,25,28^. We first assessed proliferation in pediatric patients using Ki67 expression. Pediatric COVID-19 patients had marked proliferation in the CD4 T cell compartment, with a similar frequency range of Ki67+ CD4 T cells compared to adult COVID-19 patients (**Fig S2D**). MIS-C patients were similar to pediatric COVID-19 patients in CD4 T cell proliferation (**Fig S2D**). In contrast, a higher frequency of Ki67+ CD8 T cells was observed in MIS-C compared to pediatric COVID-19 patients (**Fig S2E**). Moreover, the proliferation of CD8 T cells in MIS-C patients even exceeded that observed for the vast majority of adult COVID-19 samples (**Fig S2E**). The co-expression of HLA-DR and CD38 in humans can also be used to identify recently activated T cells likely responding to viral infection ^22,60–65,22,60–64^. Indeed, populations of HLA-DR+CD38+ CD4 and CD8 T cells were often increased in pediatric COVID-19 patients compared to HD (**Fig 2C-D**). However, MIS-C patients had substantially higher frequencies of HLA-DR+CD38+ CD4 and CD8 T cells compared to pediatric COVID-19, again rivaling or exceeding that observed in adult COVID-19 patients (**Fig 2C-D**). Among the pediatric COVID-19 patients, those with ARDS were not obviously different in T cell activation compared to non-ARDS patients, though the number of ARDS patients was low.

We next examined the distribution of T cell activation and proliferation in different CD4 and CD8 T cell subsets. In general, the patterns were consistent for pediatric COVID-19 and MIS-C cohorts and mirrored what was observed in adults. Proliferation and activation were most robust for the EM1 subset of CD8 T cells and the CM subset of CD4 T cells (**Fig S2F, S2G**). Despite the heterogeneity in the range of Ki67+ or HLA-DR+CD38+ cells, proliferation and activation were strongly correlated for individual patients and this relationship was true for both pediatric COVID-19 and MIS-C (**Fig 2E, 2F**). Together these data indicate that pediatric infection with SARS-CoV-2 resulted in robust T cell proliferation and activation. Moreover, children with MIS-C displayed marked T cell activation and proliferation, especially for CD8 T cells, even compared to severely ill adults.

In addition to conventional lymphocytes, innate and innate-like lymphocytes become activated during human viral infection and vaccination ^66–73^. In our MIS-C cohort where whole blood staining was performed, although HLA-DR expression was similar to healthy adults, more than 80% of the NK cells were CD38+ (**Fig S3A**). This increase in CD38 in MIS-C compared to pediatric COVID-19 was also observed in MAIT cells, an unconventional innate-like T cell population that also has a role in viral immunity (**Fig S3B**) ^66,74,75^. These data suggest that MIS-C is associated with activation of innate lymphocytes as well as conventional CD4 and CD8 T cells.

We next investigated the relationships between conventional lymphocyte activation and proliferation with clinical measures of inflammation and disease. In the pediatric COVID-19 cohort, there was a positive relationship between many clinical markers of inflammation and measures of T cell activation, though only the correlation between Ki67+ non-naive CD8 T cells and LDH achieved nominal significance (**Fig 2G**, top panel). In contrast, the relationships between T cell activation and clinical measures of inflammation trended in the opposite direction in MIS-C (**Fig 2G**, middle panel), although these negative associations were not statistically significant. Given the difference between groups, we next compared the directionality of the pediatric COVID-19 and MIS-C relationships. Indeed, a direct comparison demonstrated significant differences between COVID-19 versus MIS-C in how T cell activation was correlated with D-dimer or LDH (**Fig 2G**, bottom panel). These data further supported the notion that immune activation in MIS-C displays a different relationship with clinical inflammation compared to pediatric COVID-19, and together these analyses of T cell activation highlight distinct immune signatures in MIS-C.

### Distinct activation of CD8 T cell populations associated with persistent antigen and vascular surveillance in MIS-C

T cell exhaustion or dysfunction has been implicated in SARS-CoV-2 infected adults ^24,25,76^. The inhibitory receptor PD-1 is often expressed by exhausted T cells but can also be expressed by recently activated CD8 T cells ^77–81^, and in CD4 T cells PD-1 is a marker for T follicular helper cells (Tfh) ^82–85^. In adult COVID-19, PD-1 expression by CD4 T cells was elevated, reflecting changes in both CD4 T cell activation and circulating Tfh ^22^. For most pediatric patients, the frequency of PD-1+ CD4 T cells was similar to adults; however, PD-1+ CD4 T cell frequencies were substantially higher in MIS-C compared to pediatric COVID-19 (**Fig 3A**). For CD8 T cells, a similar increase in the frequency of PD-1+ cells was observed, with a substantially higher PD-1+ CD8 T cell frequency in MIS-C compared to pediatric COVID-19 (**Fig 3B**). To better assess potential CD8 T cell exhaustion versus PD-1 expression associated with recent activation, we used CD39 expression, where co-expression of PD-1 and CD39 often identifies exhausted or chronically stimulated CD8 T cells ^86–88^. Indeed, although CD39 expression by CD8 T cells alone was not significantly higher in MIS-C compared to pediatric COVID-19 (**Fig 3C**), the co-expression of both PD-1 and CD39 by CD8 T cells was substantially increased in MIS-C compared to pediatric COVID-19 (**Fig 3D**). The frequency of PD-1+CD39+ CD8 T cells in MIS-C was high even compared to severely ill adults, though there was considerable variation. The increased frequency of PD-1+CD39+ CD8 T cells in MIS-C suggests a role for prolonged antigen stimulation in the inflammatory syndrome, but the precise history or antigen exposure in the MIS-C will require further investigation.

**Figure 3.**
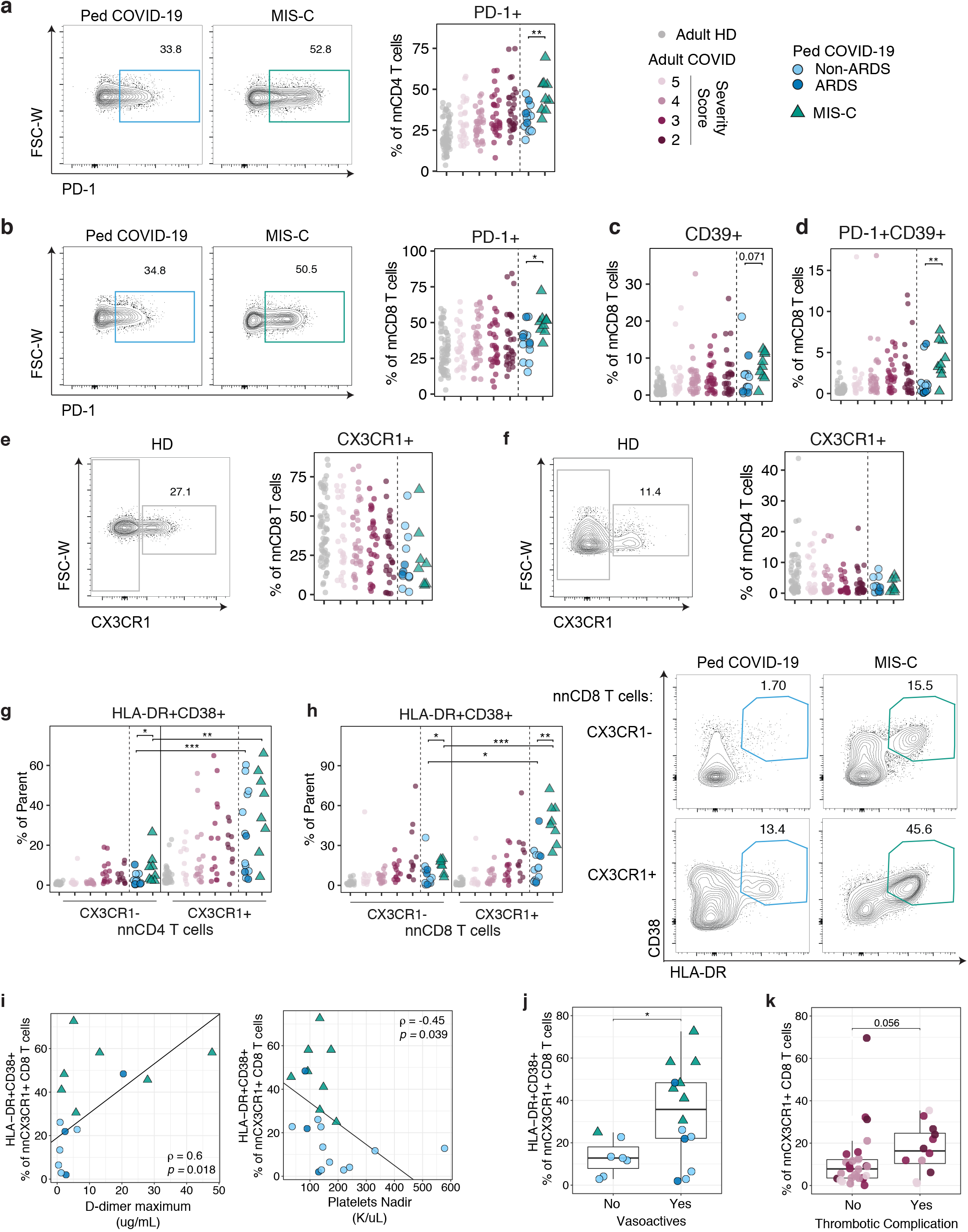
MIS-C is uniquely marked by activation in CD8 T cell populations associated with persistent antigen and vascular surveillance. (**AB**) Representative flow cytometry plots and quantification of PD-1+ nnCD4 T cells (**A**) and CD8 T cells (**B**). (**C**) Frequencies of CD39+ nnCD8 T cells. (**D**) Frequencies of CD39+PD-1+ nnCD8 T cells. (**EF**) Representative flow cytometry plots and quantification of CX3CR1+ nnCD8 (**E**) and nnCD4 T cells (**F**). (**G**) HLADR+CD38+ frequencies within CX3CR1- and CX3CR1+ nnCD4 T cells. (**H**) Representative flow cytometry plots and quantification of HLADR+CD38+ population within CX3CR1- and CX3CR1+ nnCD8 T cells. (**I**) Frequency of HLA-DR+CD38+ CX3CR1+ nnCD8 T cells versus maximum D-Dimer (left) and nadir platelet count (right). Non-parametric trend lines (Sen-Theil) for total pediatric cohort, with Spearman’s Rank Correlation coefficient (ρ) and associated P value shown. (**J**) Frequencies of HLA-DR+CD38+ CX3CR1+ nnCD8 T cells in pediatric patients categorized by treatment with vasoactive medications. (**K**) Frequencies of HLA-DR+CD38+ CX3CR1+ nnCD8 T cells in adult COVID-19 patients categorized by presence of thrombotic complication. (**A-K**) Each dot represents an individual patient or subject, with adult HD in grey circles, adult COVID-19 in shades of mauve indicated by disease severity score, Ped COVID-19 in blue circles, with ARDS patients in dark blue, and MIS-C in green triangles. (**ABCDEF**) Significance determined by unpaired Wilcoxon test between Ped COVID-19 and MIS-C groups only, indicated by: * p<0.05, ** p<0.01. (**GH**) Significance determined by unpaired Wilcoxon test between Ped COVID-19 and MIS-C groups; significance determined by paired Wilcoxon test between CX3CR1- and CX3CR1+ within pediatric groups, indicated by: * p<0.05, ** p<0.01, *** p<0.001. (**JK**) Significance determined by unpaired Wilcoxon test between clinical category, indicated by: * p<0.05.

Considerable attention has been focused on the clinical overlap with Kawasaki disease, a medium vessel arteritis of children with an unclear pathophysiology and a suggested infectious trigger ^37,41,42,89–93^. Although MIS-C and Kawasaki disease are now appreciated to be clinically distinct, both are associated with vascular involvement and shock-like presentations, though shock is less common in Kawasaki disease than in MIS-C ^37,39,40,91,94,95^. Given this vascular presentation, we investigated whether MIS-C was associated with changes in the CD8 T cell population that expresses the fractalkine receptor CX3CR1. CX3CR1+ CD8 T cells can interact with and adhere to fractalkine-expressing activated endothelium, and these interactions foster the ability of CX3CR1+ CD8 T cells to patrol the vasculature ^96–99^. The overall frequencies of CX3CR1+ CD4 and CD8 T cells were not increased in MIS-C compared to pediatric COVID (**Fig 3E, Fig 3F**). However, CX3CR1+ CD4 and CD8 T cells were both more highly activated and proliferating than CX3CR1-CD4 and CD8 T cells, based on CD38 and HLA-DR (**Fig 3G, Fig 3H**) and Ki67 expression (**Fig S4A, Fig S4B**). Moreover, compared to pediatric COVID-19, CX3CR1+ CD8 T cells in MIS-C were markedly more activated and a substantially higher proportion were Ki67+ (**Fig 3H, Fig S4B, Fig S4C**). Although CX3CR1-T cells were also more activated in MIS-C, the substantially elevated activation and proliferation of CX3CR1+ CD8 T cells in MIS-C suggested a potential role for vascular patrolling CD8 T cells in MIS-C compared to pediatric COVID-19 patients.

Although activation of CX3CR1+ CD8 T cells was highest in MIS-C, this pattern was also observed in some non-MIS-C pediatric and adult COVID-19 patients. Given these observations, and the associations made between fractalkine and vascular inflammation ^96,100–102^, we next assessed whether activated CX3CR1+ T cells correlated with vascular presentations of disease. When all pediatric patients were assessed as a group, activation of CX3CR1+ CD8 T cells was positively correlated with D-dimer levels and inversely correlated with platelet nadir (**Fig 3I**). To further investigate the potential relationship between activated CX3CR1+ CD8 T cells and vascular disease, we next asked whether the frequency of these activated CD8 T cells tracked with need for vasoactive support in pediatric SARS-CoV-2 infection. Indeed, patients requiring vasoactive medication had a considerably higher frequency of activated CX3CR1+ CD8 T cells (**Fig 3J**). This relationship was driven by MIS-C status where both the frequency of CX3CR1+ CD8 T cells and the need for vasoactive support were more common, but the association was observed in all groups (**Fig 3J**). We then asked whether these observations from the pediatric setting might inform our understanding of adult COVID-19 disease and coagulopathy. Most adults had activated CX3CR1+ CD8 T cell frequencies below those of the patients with MIS-C.

Nevertheless, the frequency of activated CX3CR1+ CD8 T cells was higher (p = 0.056) in adult COVID-19 patients with suspected or confirmed thrombotic complications (**Fig 3K**). [Note, the adult with the highest CX3CR1+ CD8 T cell activation (70% of CX3CR1+ CD8s, **Fig 3H**) had cutaneous T cell lymphoma and was undergoing active chemotherapy.] These data suggest that MIS-C is associated with distinct features of T cell activation and identify a potential novel relationship between the activation status of vascular patrolling CD8 T cells in the presentation of MIS-C and vascular complications of COVID-19 disease more broadly.

### Humoral immunity in MIS-C is dysregulated

Following infection or immunization, increased frequencies of plasmablasts (PB) can be detected transiently in the blood, peaking 1-2 weeks after initial challenge and then rapidly declining ^62,103,104^. Although the peak frequency of PB is often low, particularly following vaccination ^105–107^, in some settings of highly virulent infections PB may exceed 30% of circulating B cells^62,104,106^. Indeed, blood PB frequencies in SARS-CoV-2 infected adults can also be greater than 30% of circulating B cells ^22,25,22^, but little information exists about the responses of PB in pediatric COVID-19. We therefore evaluated the humoral response, including both antibody and cellular components of humoral immunity, in pediatric COVID-19 and MIS-C patients.

In the United States, almost all MIS-C patients are seropositive for SARS-CoV-2 specific antibodies, consistent with a delayed clinical presentation compared to acute SARS-CoV-2 infection ^40,108–110^. Both pediatric COVID-19 and MIS-C had similar frequencies of naive B cells, CD27-IgD-B cells, non-switched memory, and switched memory B cells (**Fig S5A-C**). However, given the likely delayed clinical presentation of MIS-C relative to SARS-CoV2 infection, we hypothesized that MIS-C patients would be past the peak PB response and would therefore have lower PB frequencies compared to pediatric COVID-19 patients. Instead, PB frequencies were elevated in both pediatric COVID-19 and MIS-C compared to healthy adults, suggesting ongoing humoral responses in both settings (**Fig 4A**). As observed in adults ^22^, the blood PB frequency did not correlate with spike receptor binding domain (S-RBD)-specific IgM or IgG in either cohort (**Fig 4B**). Given the inflammatory environment of COVID-19 and MIS-C, we next assessed correlations with clinically measured plasma cytokines. PB frequencies were nominally correlated with plasma IFNγ, and naive B cells were inversely correlated with IFNγ (**Fig S5D**)^111–113^. Furthermore, a shift from naive B cells towards CD27-IgD-was associated with IL-6. In contrast, there were no significant associations between activated or proliferating T cell subsets and cytokines (**Fig S5E**). Together these data suggest that the PB responses are elevated in MIS-C and that B cell responses maintain a similar relationship to the measured plasma cytokines as is observed in pediatric COVID-19.

**Figure 4.**
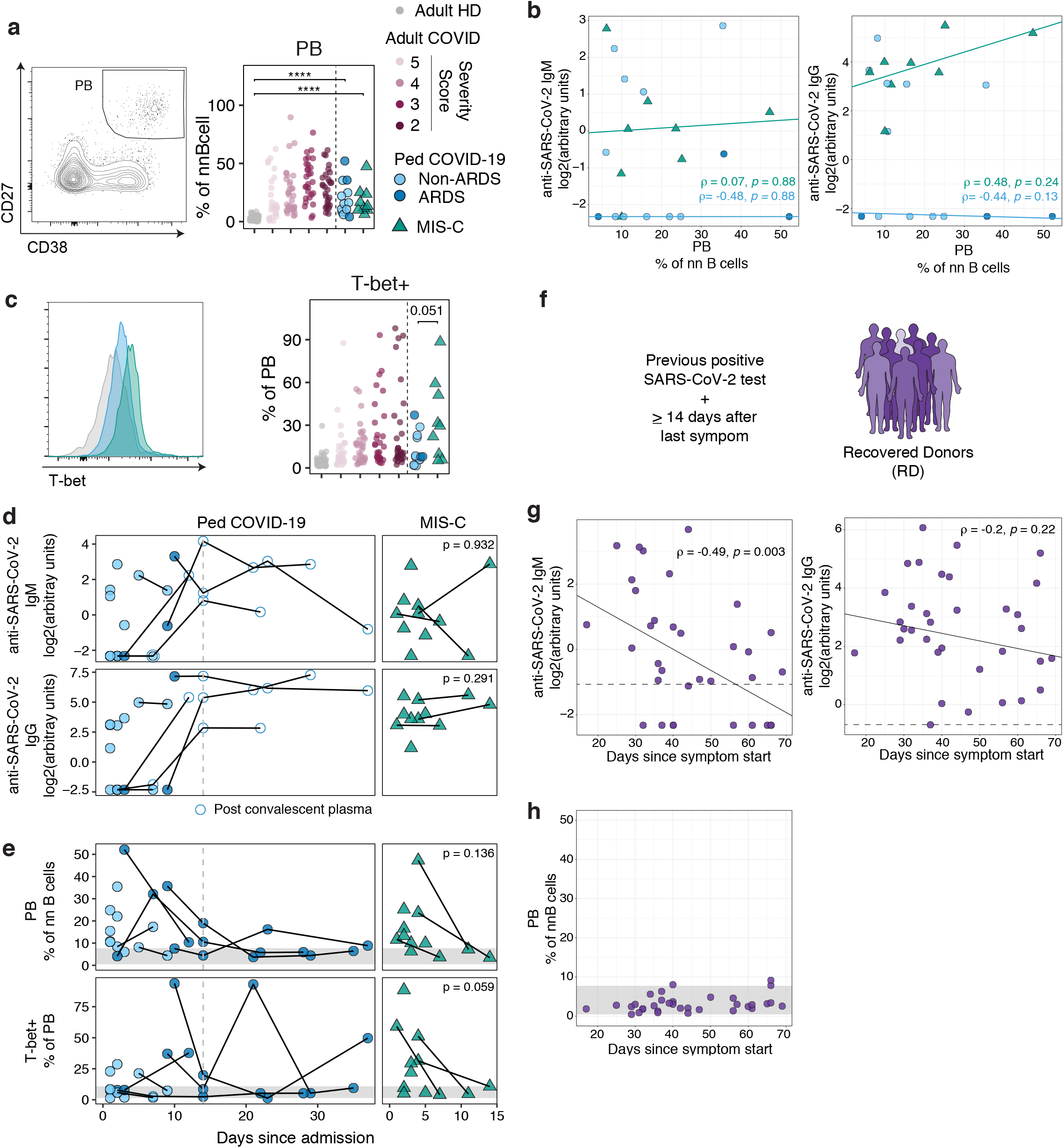
Humoral immunity in MIS-C is dysregulated. (**A**) Representative flow cytometry plots and quantification of plasmablasts (PB). For comparison between pediatric cohorts and adult HD, significance was determined by unpaired Wilcoxon test, indicated by **** p<0.0001. (**B**) Frequency of PB within nnB cells versus anti-SARS-CoV-2 IgM (left) and IgG (right). Non-parametric trend lines (Sen-Theil) for each pediatric cohort, with Spearman’s Rank Correlation coefficient (ρ) and associated P value shown. (**C**) Representative histogram of T-bet expression by PB for an adult HD (grey), Ped COVID-19 (blue) and MIS-C (green) and quantification of frequencies of T-bet+ PB. Significance determined by unpaired Wilcoxon test between pediatric cohorts. (**D**) Anti-SARS-Cov-2 antibody levels and (**E**) B cell features over days since admission in Ped COVID-19 (left) and MIS-C subjects (right). Black lines connect repeat draws for individual subjects; for MIS-C, paired t-test P value is shown for the three subjects with repeat draws. (**F**) Overview of adult recovered donor (RD) cohort. (**G**) Anti-SARS-CoV-2 IgM (left) and IgG (right) versus days since symptom start for RD cohort. Non-parametric trend lines (Sen-Theil) with Spearman’s Rank Correlation coefficient (ρ) and associated P value shown. Dotted grey line denotes positive threshold. (**H**) Frequency of PB within nnB cells versus days since symptom start for RD cohort. Grey shading indicates the derived central 95% adult HD reference interval. (**A-E**,**G-H**) Each dot represents an individual patient or subject, with adult HD in grey circles, adult COVID-19 in shades of mauve indicated by disease severity score, RD in purple circles, Ped COVID-19 in blue circles, with ARDS patients in dark blue, and MIS-C in green triangles. (**EH**) Grey shading indicates the derived central 95% adult HD reference interval.

Although total PB frequencies were comparable between pediatric COVID-19 and MIS-C, the differentiation state of the PB was distinct. Specifically, the T-box transcription factor T-bet was higher (p=0.051) in PB from MIS-C compared to pediatric COVID-19, with similar observations for the T-box transcription factor Eomesodermin (**Fig 4C, S5F**). T-bet expression in B cells has been associated with extra-follicular responses, advanced age, autoimmunity, and viral infections where T-bet has a role in class switching to antiviral isotypes ^113–116^. These data suggest that, although similar in frequency, the differentiation state of PB responding in MIS-C patients is altered compared to pediatric COVID-19.

We next assessed the T cell component of humoral immunity, Tfh. Like PB, circulating Tfh that express ICOS and/or CD38 (activated cTfh) increase in blood 1-2 weeks after immunologic challenge ^107,117–120^. Activated cTfh enrich for the transcriptional and epigenetic signatures of Tfh in germinal centers and can provide insights into the coordination of the humoral response to infection or vaccination ^120,121^. As expected from studies in influenza vaccination and adult COVID-19 ^22,117^, total cTfh frequencies were similar between pediatric COVID-19, MIS-C, and healthy adult donors (**Fig S6A**). The frequency of activated cTfh was also similar to healthy adult donors (**Fig S6B**). Moreover, total or activated cTfh populations did not correlate with the PB response in either pediatric cohort (**Fig S6C-E**). Of note, both the frequency and gMFI of CXCR5 were reduced in pediatric COVID-19 (gMFI only) and MIS-C compared to healthy adults (**Fig S6F**). The lower expression of this follicular-homing chemokine receptor could impact coordination of the germinal center and humoral response or be a symptom of follicular disruption.

We next examined the stability of S-RBD-specific IgM and IgG as well as PB in the eight patients who underwent repeated research blood draws (5 COVID-19, 3 MIS-C). In both pediatric COVID-19 and MIS-C, the level of spike receptor binding domain (S-RBD)-specific IgG and IgM did not change substantially over the first 15 days of admission, unless patients received convalescent plasma (open circles, **Fig 4D**)^122^. Due to different clinical courses, pediatric COVID-19 patients and patients with MIS-C did not have the same availability for additional research blood draws. While patients with ARDS remained hospitalized for weeks, the median duration of hospitalization in MIS-C was 8 days. Patients with ARDS had stable antibody responses into weeks three and four of hospitalization, although it is unclear whether these humoral responses were due to induction of host immunoglobulin production or reflected antibody from the convalescent plasma, as we previously reported ^122^. In MIS-C, the frequencies of both total PB and Tbet+ PB consistently decreased during hospitalization, with PB responses similar to healthy adults by the second draw in every case (**Fig 4E**). In pediatric COVID-19, PB changes were more varied. Although PB frequencies decreased to the HD range in many cases, in two individuals these frequencies increased during the first 15 days of admission. A similar pattern was observed for Tbet+ PB. Together these data suggest that MIS-C is associated with stable antibody responses over time but with declining PB frequency and a change in PB differentiation state during clinical improvement.

The elevated PB frequencies in MIS-C are perhaps surprising, given the hypothesis that MIS-C is a delayed clinical presentation of a SARS-CoV-2 infection occurring several weeks earlier. One possibility is that SARS-CoV-2 infection typically results in a prolonged PB response in the blood. To further investigate this possibility, we examined the duration of elevated PB frequencies in our previously published cohort of recovered adult donors (RD). RD were symptomatic individuals who were diagnosed with SARS-CoV-2 infection by PCR, never hospitalized, and then experienced resolution of all symptoms (**Fig 4F**)^22^. For these subjects, we collected the date of symptom start and were therefore able to plot a cross-sectional analysis of individual measures of humoral responses relative to self-reported symptomatic disease. Over the course of 17-69 days since symptom initiation, the quantity of S-RBD-specific IgM decreased, whereas S-RBD-specific IgG was detectable in every RD and remained relatively stable over time (**Fig 4G**). However, PB frequencies in RD were similar to adult healthy donor PB frequencies (grey bar), even within the first month after symptom start (**Fig 4H**), suggesting rapid resolution of the PB response in recovered adult COVID-19 patients. The high PB frequencies at clinical presentation for MIS-C, together with the notion that MIS-C clinical presentation occurs at least 3-4 weeks post infection ^35,110,123,124^ suggest a scenario where either the initiation of the adaptive immune response in MIS-C is delayed relative to infection and peaks 3-4 weeks post infection or is prolonged, remaining detectable upon hospitalization. In either case, patients with MIS-C resolved these PB features as they clinically improved.

### Immune perturbations in MIS-C overlap with severe adult COVID-19 and correct coincident with clinical improvement

We next sought to understand the global immune landscape of pediatric SARS-CoV-2 infection and in particular MIS-C. To ask in an unbiased manner whether the pediatric immune landscape was distinct from COVID-19 adults, we built a Uniform Manifold Approximation and Projection (UMAP) to distill 207 immune features into two-dimensional space. In comparison to our previous study of only adults ^22^, here the data used to generate the UMAP included pediatric patients in addition to the healthy and COVID-19 adults. Furthermore, as in **Fig 2** and **3**, features included in this UMAP were restricted to non-naive T and B cells to eliminate the strongest source of age-associated bias. Consistent with the prior model ^22^, adult HD and COVID-19 adults were immunologically distinct (**Fig 5A**). Notably, the immune landscape of pediatric MIS-C and most pediatric COVID-19 patients was generally located with COVID-19 adults rather than adults who were healthy. MIS-C patients, in particular, more consistently overlapped with adults who were severely ill.

**Figure 5.**
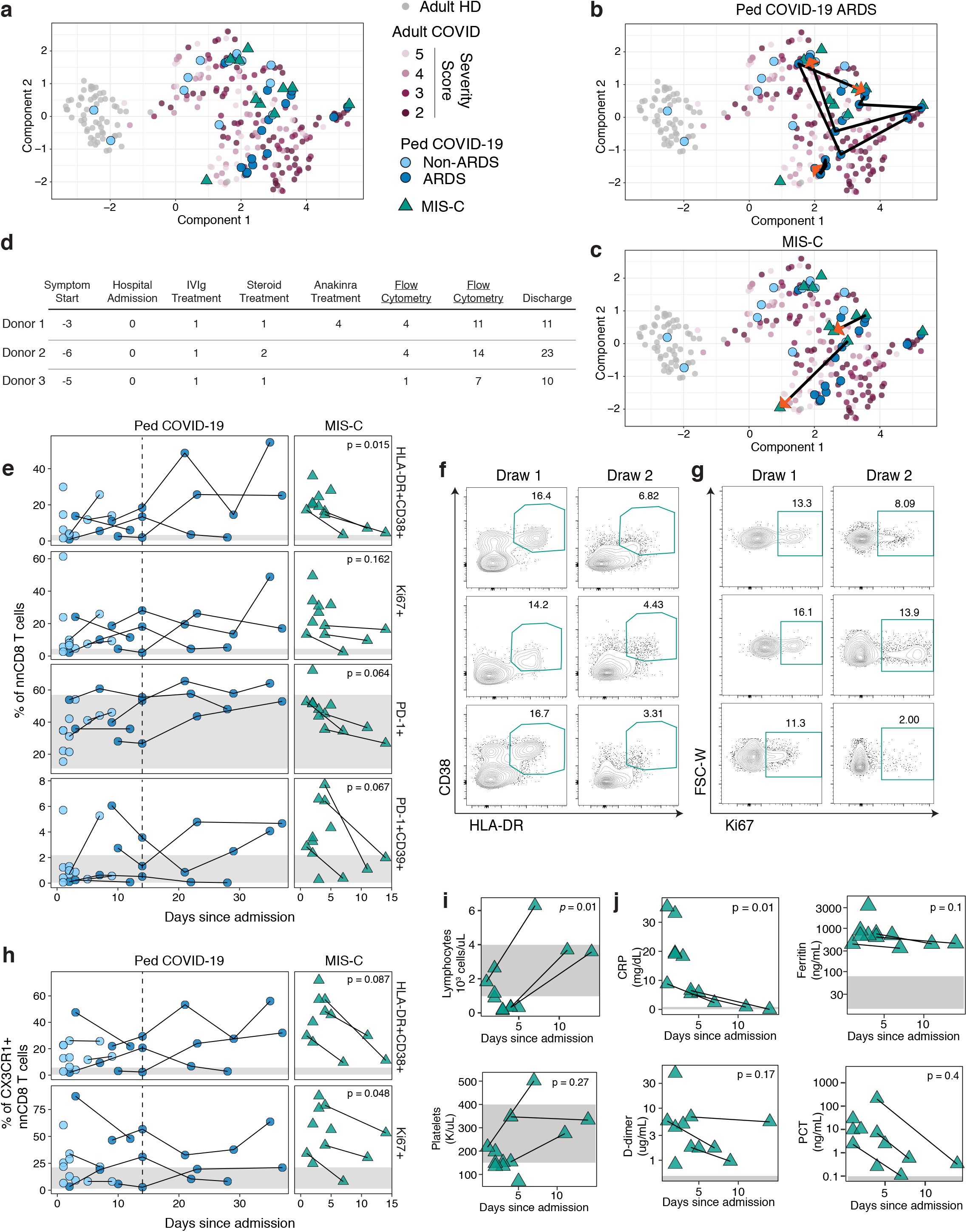
Immune perturbations in MIS-C overlap with severe adult COVID-19 and correct coincident with clinical improvement. (**A**) Transformed UMAP projection of aggregated flow cytometry data from PBMC analysis. (**B**) Trajectory in UMAP space for repeat Ped COVID-19 ARDS draws. (**C**) Trajectory in UMAP space for repeat MIS-C draws. (**D**) Clinical course for MIS-C subjects where repeat samples were available. (**E**) Activated and proliferating CD8 T cell populations over days since admission in Ped COVID-19 (left) and MIS-C subjects (right). (**F**) Flow cytometry plots for HLA-DR+CD38+ nnCD8 for MIS-C patients with repeat draws. (**G**) Flow cytometry plots for Ki67+ nnCD8 for MIS-C patients, as in (**F**). (**H**) CX3CR1+ HLA-DR+CD38+ and Ki67+ CD8 T cell populations over days since admission in Ped COVID-19 (left) and MIS-C subjects (right). (**I**) Lymphocyte and platelet counts over days since admission for MIS-C subjects. (**J**) Clinical inflammatory markers over days since admission for MIS-C subjects. (**A-C**,**E**,**H-J**) Each dot represents an individual patient or subject, with adult HD in grey circles, adult COVID-19 in shades of mauve indicated by disease severity score, Ped COVID-19 in blue circles, with ARDS patients in dark blue, and MIS-C in green triangles. (**EHIJ**) Black lines connect repeat draws for individual subjects. For MIS-C, paired t-test test P value is shown for the three subjects with repeat draws. (**EH**) Grey shading indicates the derived central 95% adult HD reference interval. (**IJ**) Grey shading indicates normal clinical laboratory reference ranges for pediatric subjects.

Given differences in clinical trajectory between patients admitted to the pediatric ICU with ARDS versus MIS-C, we next investigated whether the clinical course was mirrored in the UMAP high dimensional immunologic space. We first asked how the immune landscape of the patients with ARDS changed over the course of hospitalization (**Fig 5B**). One patient with ARDS demonstrated a stable immunophenotype over time, with all 5 timepoints in close proximity on the UMAP, whereas two other patients displayed considerable movement in UMAP space indicating temporal changes in the overall immune response. We next assessed the trajectory for 2 of the 3 MIS-C patients on whom repeat draws and high quality flow cytometric data existed for all UMAP features (**Fig 5C**). Both MIS-C patients moved towards a position enriched for less ill adults (NIH score 4-5) and towards HD. The two MIS-C patients were 0 and 3 days from hospital discharge at the second research blood draw (donor 1 and donor 3, **Fig 5D**). Thus, at least for the patients with multiple time points available, MIS-C was associated with global immune landscape changes in the UMAP towards locations associated with less severe disease in adults.

We next asked whether the MIS-C position changes on the UMAP were associated with decreased T cell activation as patients progressed towards hospital discharge. In ARDS patients, CD4 T cells initially displayed minimal activation and were similar to healthy adults for HLA-DR, CD38, Ki67 as well as PD-1 and CXCR5 expression (**Fig S7A-C**). In contrast, MIS-C patients initially demonstrated more CD4 T cell activation than healthy adults by most metrics, but this activation resolved over time (**Fig S7A-C**). Similarly, the high PD-1 expression and low CXCR5 expression initially observed for CD4 T cells in many MIS-C patients was also resolved during the second week of hospitalization.

Unlike CD4 T cells, many features of CD8 T cell activation and proliferation were elevated compared to the healthy adult range at admission for pediatric COVID-19 and MIS-C patients (**Fig 5E**). One exception was PD-1, where the range of expression for healthy adults was wide. In patients with prolonged admission due to ARDS, however, CD8 T cell activation and proliferation remained elevated over time (**Fig 5E**). This pattern contrasted with MIS-C patients, where HLA-DR+CD38+ CD8 T cells decreased in the first two weeks of admission (**Fig 5E, 5F**) and PD-1+CD39+ CD8 T cells fell to the range observed in healthy adults (**Fig 3D, Fig 5E**). The frequency of Ki67+ CD8 T cells in MIS-C also decreased in the second week of admission, though proliferating CD8 T cells remained elevated compared to healthy adults for two of three patients (**Fig 5E**,**5G**). Similarly, proliferating or activated CX3CR1+ CD8 T cells decreased substantially as MIS-C patients moved towards hospital discharge, but remained above the range in healthy adults (**Fig 5H**). In contrast, in ARDS patients where activation or proliferation of CX3CR1+ CD8 T cells was above the normal range, these CD8 T cells remained activated over time (**Fig 5H**). Thus, the different temporal patterns of CD8 T cell activation may explain continued co-localization of ARDS with severely ill adults in the UMAP analysis, but movement of the MIS-C patients towards UMAP locations associated with less severe disease over time.

The concept that children with MIS-C resolve some but not all of their immunologic perturbations at hospital discharge was also reflected in many standard clinical laboratory measurements. The immunophenotypic changes corresponded to correcting lymphocyte and platelet counts, although in one patient the absolute lymphocyte and platelet counts rose above the normal range towards the end of the hospitalization (**Fig 5I**). Elevated CRP, D-dimer, PCT and ferritin all declined over time in MIS-C (**Fig 5J**), although only CRP declined to the normal range by the second research blood draw. Therefore, these clinical measures in concert with cellular measures of immune dysregulation suggest an ongoing resolution of pathology, though this resolution remains incomplete near the time of discharge. Together these data reveal a profound immunologic perturbation in MIS-C that, although overlapping with features of adult patients and pediatric patients with ARDS at clinical presentation, appear to begin resolving immunologically coincident with clinical improvement.

## DISCUSSION

The pathogenesis of SARS-CoV-2-associated MIS-C remains poorly understood. This unusual presentation of SARS-CoV-2 infection is likely immunologically mediated as MIS-C patients demonstrate markedly elevated clinical measures of inflammation and respond to immune modulation with IVIg and steroids. The underlying immune landscape of MIS-C, including temporal changes in immune responses, has remained underexplored. Here, to address these questions and to interrogate immune dysregulation in MIS-C versus pediatric COVID-19, we performed deep immunologic profiling of 14 children with MIS-C and 16 children with COVID-19. We compared innate and adaptive cellular immunity in children to healthy and COVID-19 hospitalized adults. Our studies revealed distinct features of lymphocyte activation and humoral immunity that give insights into MIS-C pathogenesis and immunologic shifts over time in pediatric COVID-19.

One of the more striking differences between MIS-C and COVID-19 was activation of CX3CR1+ CD8 T cells. CX3CR1 is a chemokine receptor expressed by microglia, myeloid cells and lymphocytes that binds the ligand CX3CL1 ^125,126^. For lymphocytes, CX3CR1 expression is highly biased to cytotoxic, effector-like CD8 T cells and NK cells and, in particular, expressed by a CD8 T cell population that patrols vasculature with a proposed role in control of persisting and/or reactivating viral infection ^96,98,127^. Moreover, this CX3CR1-CX3CL1 axis has a key role in cardiovascular disease, where CX3CL1 is expressed and presented by vascular endothelial cells to mediate adhesion and then subsequent extravasation of leukocytes ^128^. CX3CL1 expression can be increased by inflammatory cytokines present during viral infections including TNF, IFN-γ and IL-1 and, in cardiovascular disease, this increase in chemokine in the vasculature leads to increased recruitment of CX3CR1+ T cells ^128^. Thus, the identification of a major difference in activation of CX3CR1+ CD8 T cells in MIS-C may have direct relevance to the vascular pathology observed in these patients. Indeed, this immunological phenotype was associated with a requirement for vasoactive support, elevated D-dimer, and decreased platelets in pediatric patients. Further, activated CX3CR1+ CD8 T cells decreased coincident with clinical improvement. It will be interesting in the future to assess whether there is a causal relationship between changes in activation of these CX3CR1+ CD8 T cells vascular disease presentation and/or resolution.

The identification of CX3CR1+ CD8 T cell activation was possible because of the dichotomous classification of pediatric COVID-19 versus MIS-C presentations. However, these data also point to potential mechanisms of disease in adults. Several studies suggest that severe COVID-19 in adults is, in part, a vascular disease ^129–132^, and a MIS-C like presentation has also been observed in adults ^33,133^. Although the age of patients with MIS-C (median of 8 years^40^) likely provides a clue to MIS-C pathogenesis, it is possible that similar mechanisms of disease can occur across the age spectrum. In particular, our studies identify a potential relationship between thrombotic complications in adult COVID-19 patients and activation of CX3CR1+ CD8 T cells. Although these data suggest a possible common feature of these SARS-CoV-2 illnesses, it should be noted that the definition of thrombotic complications in adults is substantially more broad than for MIS-C, highlighting the need for further studies investigating these potential relationships. Nevertheless, inhibitors of CX3CR1 exist, have been used in preclinical models ^134^, and could be interesting options for clinical development. Moreover, genetic polymorphisms in *CX3CR1* are associated with risk of coronary artery disease ^135^, further highlighting the potential role of this pathway in immune connections to vascular disease, perhaps providing insights for MIS-C and adult cardiovascular complications of COVID-19. It will be interesting in the future to investigate potential genetic associations with cardiovascular risks in COVID-19 patients and these genetic variants. Moreover, these observations may provide additional insights into other infection-related vascular complications including Kawasaki disease.

The current studies also highlight the potential temporal differences in humoral immunity in MIS-C, compared to acute pediatric COVID-19 and resolved adult disease. In US cohorts, patients with MIS-C are almost universally seropositive, unlike pediatric and adult presentations of COVID-19 ^40,108–110^. Seropositivity is consistent with the notion that MIS-C is a delayed event, presenting weeks after initial SARS-CoV-2 infection (i.e. with enough time for antiviral antibody to develop). The substantially elevated PB frequencies we observed in MIS-C are, therefore, perhaps surprising. In adults who recover from COVID-19, PB frequencies return to baseline 2-4 weeks after symptoms resolve, whereas the PB responses in a subset of adult COVID-19 patients who remain hospitalized can be prolonged ^22^. The reasons for increased PB frequency in MIS-C at admission are unclear but could reflect either aberrantly sustained PB production or a newly initiated response. The presence of a higher proportion of T-bet+ PB in MIS-C could be consistent with an aberrant or extrafollicular response ^113,115,116^, but in other settings T-bet expression in B cells is required to control viral infection and aids in the development of antibody secreting cells ^136–138^. One study of MIS-C suggested altered quality of humoral responses with low antibody breadth and decreased antibody neutralizing activity ^109^; however, increased T-bet expression in our study was not accompanied by altered antibody breadth or neutralization in this cohort when compared to pediatric COVID-19 ^108^. Of note, PB and T-bet+ PB frequencies in MIS-C declined rapidly after hospitalization. This decrease could reflect the idea of a temporally more advanced immune response in MIS-C that resolves with time or reflect a response to immune modulation with IVIG and/or steroids. Together our data support skewed B cell responses in MIS-C, but future studies will be necessary to dissect whether and how germinal center reactions are altered in this novel inflammatory syndrome.

Our findings provide several key insights into the potential drivers of immune pathogenesis in MIS-C. One possibility suggested by our data is continued activation of adaptive immune responses, driven by persisting antigen. In this context, MIS-C may reflect the latter stage of a poorly controlled primary infection. Indeed, CD8 T cell activation, elevated frequencies of PD-1+CD39+ CD8 T cells, and robust PB in the blood at clinical presentation in MIS-C are all consistent with prior reports of responses to persisting antigen ^86–88^. The potential role for T cell exhaustion in COVID-19 remains poorly understood and it is often challenging to distinguish true exhaustion from recent T cell activation. However, the long pre-symptomatic phase and rapid development of severe MIS-C may be inconsistent with a chronic or indolent infection. Rather, these clinical observations suggest a second possibility: an additional trigger occurring ∼2-3 weeks after the initial infection with SARS-CoV-2. This new event could either be virologic (e.g. SARS-CoV-2 localizes to a new tissue type), due to a secondary or perhaps pre-existing infectious trigger, or auto-reactive (e.g. prolonged asymptomatic infection or inflammation ignites auto-reactive responses). Whatever the cause, one consequence is marked lymphocyte activation, and particularly activation of CX3CR1+ vascular patrolling CD8+ cells. Future studies will be necessary to investigate the clonality and specificity of activated lymphocytes in MIS-C compared to COVID-19, to identify the targeted SARS-CoV-2 antigens, and to determine the potential contribution of bystander activation, homeostatic proliferation following lymphopenia, or auto-reactive cells ^29,139–142^. Third, our observation of a sustained PB response with increased expression of T-bet highlights a potential role for antibody in MIS-C pathogenesis. Further studies are required to assess whether such sustained PB responses are SARS-CoV-2 specific or an auto-reactive antibody response. However, dysregulated and/or auto-reactive antibody responses could be linked to the observed disrupted germinal center architecture in SARS-CoV-2 infection, perhaps through increased extrafollicular activity ^115,143,144^. Taken together, studies of antigen specificity will be critical in understanding lymphocyte activation and immune dysregulation in MIS-C.

In summary, deep immune profiling of >200 immune parameters measured in the peripheral blood of pediatric COVID-19 and MIS-C reveal similarities of pediatric COVID-19 patients with adult COVID-19 patients in many respects. However, the activation profiles of MIS-C often aligned more strongly to adults with more moderate-to-severe disease, suggesting a more robust response, with these immune activation patterns resolving over time. In contrast, severely ill ARDS pediatric COVID-19 patients maintained immune activation features and aligned with features of severely ill adults. Future studies that integrate our findings with immunologic profiles from both children and adults with a wide spectrum of disease, including asymptomatic patients, should further define the heterogeneity of immune responses and the role of immune pathogenesis in SARS-CoV-2 infection. Comparing and contrasting the immune system in distinct clinical presentations of SARS-CoV-2 infection will help guide precision immunotherapeutics in children and may shed light on the pathology of severe disease in diverse COVID-19 patient populations.

## Data Availability

The authors confirm that the data supporting the findings of this study are available within the article and its supplementary materials.

## Acknowledgements

The authors thank patients and blood donors, their families and surrogates, and medical personnel. We additionally thank Daniel Corwin and Fran Balamuth for maintenance of an extensive literature review of the early COVID-19 clinical science.

## Author Contributions

LAV, JRG, AEB, SaEH, and EJW conceived the project. LAV, JRG, AEB, DAO, and EJW designed the study. CD, EMB, HB, SaEH, and DTT conceived the pediatric clinical cohorts. CD, KOM, and JC enrolled subjects and coordinated sample collection. AEB and KDA coordinated sample processing. AEB, LKC, MBP, JEW, ZC, YJH, CB, CD, and DM performed experiments. JHL, CJ, CD, KOM, JC, HB, HMG, and DTT abstracted clinical metadata. DAO and CA integrated pediatric and adult clinical and serologic data. NJM conceived the adult COVID-19 cohort. LAV, EJW, and OK conceived the healthy and recovered adult cohorts. DAO performed data validation. JRG, AEB, DAO, LAV, LKC, and MBP analyzed data. EMA, SG, and ScEH performed serologic studies. LAV, JRG, AEB, DAO, and EJW wrote the manuscript. All authors reviewed the manuscript.

## MATERIALS AND METHODS

### Human Subjects

#### Pediatric subjects

Peripheral blood samples were collected from patients ages 18 and younger admitted to the Children’s Hospital of Philadelphia with evidence of current or past SARS-CoV-2 infection by PCR or serologic testing. MIS-C was defined as presentation with fever, clinically severe illness, and multisystem organ involvement (>2 of cardiac, renal, respiratory, hematologic, gastrointestinal, dermatologic, or neurologic), along with positive SARS-CoV-2 PCR or serology. Patients with suspected MIS-C could be enrolled before SARS-CoV-2 serology results were received. Patients with COVID-19 were PCR positive and included patients with a clinical diagnosis of acute respiratory distress syndrome (ARDS). COVID-19 patients without ARDS broadly included those admitted for SARS-CoV-2 associated disease as well as patients found to have SARS-CoV-2 incidentally. Some patients in this cohort are described in prior publications ^32,36,108,122^. The research protocol was approved by the CHOP Institutional Review Board. Due to the COVID-19 pandemic, verbal informed consent was obtained from a legally authorized representative as per the Declaration of Helsinki. Written informed consent was signed by the consenting physician and a copy was provided to participants. After consent was obtained, peripheral blood was collected. Repeat samples were obtained in a subset of patients who remained in the hospital. <u>Adult subjects</u>: Peripheral blood samples from adults were collected as previously described ^22,25^. Briefly, patients admitted to the Hospital of the University of Pennsylvania with a positive SARS-CoV-2 PCR were screened and approached for informed consent. COVID-19 subjects were classified according to an ordinal scale: (1) death, (2) hospitalized, on invasive mechanical ventilation or ECMO or both, (3) hospitalized, requiring nasal high flow oxygen therapy, non-invasive mechanical ventilation, or both, (4) hospitalized, requiring supplemental oxygen, (5) hospitalized, not requiring supplemental oxygen or on pre-hospital baseline oxygen, (6) non-hospitalized, but unable to resume normal activities, (7) non-hospitalized, with resumption of normal activities. Healthy donors (HD) were adults with no prior diagnosis of or recent symptoms consistent with COVID-19. Recovered COVID-19 subjects (RD) were adults with a prior positive SARS-CoV-2 PCR test by self-report who met the definition of recovery by the Centers for Disease Control.

### Clinical data abstraction

For inpatients, clinical data were abstracted from the electronic medical record into REDCap databases. Clinical laboratory data were abstracted as maximum, minimum, or from the date closest to research blood collection (e.g. 1st research blood draw, **Supplementary Table 1**). Co-infections were identified in 6 patients (**Supplemental Table 1** and as previously described ^32^). Obesity was defined as body mass index > 95%ile. HD and RD completed a survey about symptoms. Adult subjects were categorized by the clinical team as having suspected or confirmed thrombotic complications, defined as deep venous thrombosis, pulmonary embolism, arterial thrombus, myocardial infarction (with or without ST elevation), cerebrovascular accident, or continuous hemodialysis circuit clotting. Given low numbers of each individual complication, these thrombotic complications were considered in aggregate, and some subjects had more than one. Diagnosis was based on clinical suspicion, and there was no routine screening employed beyond baseline electrocardiogram on ICU admission. All participants or their surrogates provided informed consent in accordance with protocols approved by the regional ethical research boards and the Declaration of Helsinki.

### Blood Processing

Peripheral blood was collected into sodium heparin tubes (BD, Cat#367874) and processed for staining from whole blood or for PBMC, as previously described ^22,25^. Briefly, for whole blood leukocyte staining, an aliquot of whole blood was removed and treated with ACK lysis buffer (ThermoFisher) to remove red blood cells. To isolate PBMC, remaining whole blood was spun, plasma removed, and the blood pack was resuspended in RPMI. PBMC were isolated using SEPMATE tubes (STEMCELL Technologies, Cat#85450), following the manufacturer’s protocol. PBMC were then washed, treated with ACK lysis buffer (ThermoFisher), and filtered with a 70uM filter before flow cytometric staining.

### Antibody panels and staining

PBMC were stained as previously described<u>(Mathew et al. 2020)</u>. See **Supplementary Table 2** for antibody panel information. Briefly PBMC were stained with a live/dead stain before treatment with Fc block. All chemokine receptors were stained for 20 min at 37°C. Additional surface stain and secondary antibody stain was performed at RT. PBMC were fixed and permeabilized, then stained for intracellular markers at 4°C overnight. The following day, samples were fixed in 4% PFA. Samples were acquired immediately in 1% PFA. Whole blood cell staining was performed as described previously<u>(Kuri-Cervantes et al. 2020)</u>. See **Supplementary Table 3** for antibody panel information. Briefly, chemokine receptors were stained for 10 min at 37°C, followed by staining with a viability dye and surface markers at RT. Samples were fixed, permeabilized and stained for intracellular markers at 37°C for 1 hour, before a final fix in 4% PFA.

### Flow Cytometry

Samples were acquired on a 5 laser FACS Symphony A5 (BD Biosciences), as described previously ^22,25^. Optimized photomultiplier tube voltages (PMTs) were determined by voltration and tracked over time using SPHERO rainbow beads (Spherotech, Cat#RFP-30-5A). Compensation was performed using UltraComp eBeads for PBMC study (ThermoFisher, Cat#01-2222-42) or BD CompBeads (BD Biosciences, Cat#552843 and 552844). Up to 2⨯10^6^ live PBMC or 5⨯10^6^ total events from whole blood were acquired per each sample.

### Flow Cytometric Analyses

All analysis and visualization was performed in FlowJo (Treestar, version 10.6.2). Major lymphocyte populations were identified as shown in **Figure 1H**. Of note, compared to the strategy used in Mathews et al., an additional gate was added to exclude CD16^bright^ SSC-A^high^ monocyte-like cells. As previously described, batch correction was performed for samples acquired before and after a panel change to remove one antibody. Briefly, a variance stabilizing transform was applied to each of the primary flow features separately (logit for fraction-of-parent features and logarithm for gMFI features), the mean of the second panel was centered to match the first, and the result was inverse-transformed back to the original scale.

### Serology

Enzyme-linked Immunosorbent Assays (ELISAs) were performed using plates coated with the receptor binding domain of the SARS-CoV-2 spike protein, as previously described ^108,145^. Briefly, plasma samples were diluted 1:50 for initial assays. If the IgG or IgM concentration was above the lower limit of detection (positive, set at 0.20 arbitrary units), the sample was then run in at least a 7-point dilution series for quantitation. A dilution series of the monoclonal antibody IgG CR3022 (specific for the SARS-CoV-2 spike protein) was used as a control across plates.

### Plasma Cytokine Analyses

Blood collected into lithium heparin tubes was spun for plasma isolation in pediatric subjects. Cytokine profiling was performed using V-Plex Pro-inflammatory Panel 1 Human Kits (Cat. #K15049D; Meso Scale Diagnostics, Rockville MD, USA). Samples were tested in duplicate and results measured using the QuickPlex SQ120 (Meso Scale Diagnostics).

### Statistics

Unless otherwise noted, non-parametric tests were used throughout this study to accommodate for the heterogeneity of clinical and research data. Correlation was quantified by Spearman’s rank correlation coefficient (ρ). Associations between ordered features (e.g. continuous features measured by flow cytometry or a clinical laboratory value like D-dimer) were evaluated by Spearman’s rank correlation test. Associations between discrete unordered features (e.g. patient sex or binary encoding of comorbidities such as obesity) were evaluated by Fisher’s exact test. Associations between mixed ordered versus unordered features were performed by unpaired Wilcoxon test. Discrete unordered features with > 2 categories (e.g. ABO blood type) were expanded into binary “dummy” variables prior to testing by the methods described immediately above. To test for a difference in the Spearman rank correlations between COVID-19 and MIS-C for specified features in Figure 2G, a non-parametric permutation test was used. Briefly, all underlying paired data were held fixed and a difference statistic, Δρ = ρ_MISC_ - ρ_COVID_, was computed under the exact null distribution derived by all unique permutations of the binary COVID-19 versus MIS-C label. Trajectory analysis was performed by paired T-test across all MIS-C patients with paired sequential blood draws. All other paired analyses were performed by non-parametric paired Wilcoxon test. All tests were performed two-sided with a nominal significance threshold of P < 0.05. Where appropriate, multiple test correction was performed using the Benjamini-Hochberg procedure at the FDR < 0.05 significance threshold. Other details are provided within the relevant figure legends.

