## Supplementary Materials for "Deep Immune Profiling of MIS-C demonstrates marked but transient immune activation compared to adult and pediatric COVID-19"

for


Supplementary Figure Legends

Supplemental Table 1

Supplemental Table 2

Supplemental Table 3

Supplemental Figure 1

Supplemental Figure 2

Supplemental Figure 3

Supplemental Figure 4

Supplemental Figure 5

Supplemental Figure 6

Supplemental Figure 7

#### SUPPLEMENTAL FIGURE LEGENDS

**Supplemental Figure 1 - Innate immune compartment in MIS-C is broadly comparable to Pediatric COVID-19**

(**A**) White blood cell (WBC) counts in pediatric cohorts at time of draw. (**B**) Clinically determined cell counts in pediatric cohorts at time of draw. (**C**) Gating strategy and representative flow cytometry plots for immune populations from whole blood stain. (**D**) Frequencies of eosinophils, neutrophils and immature granulocytes across all cohorts. (**E**) Monocyte and subset frequencies across all cohorts. (**F**) Dendritic cell (DC) and subset frequencies across all cohorts. (**ABDEF**) Each dot represents an individual patient or subject, with adult HD in grey circles, adult COVID-19 in shades of mauve indicated by disease severity score, Ped COVID-19 in blue circles, with ARDS patients in dark blue, and MIS-C in green triangles. (**AB**) Normal clinical laboratory reference ranges for healthy pediatric subjects are indicated in grey shading. (**DFE**) Significance determined by unpaired Wilcoxon test between Ped COVID-19 and MIS-C groups only, indicated by: * $p<0.05$, or P value shown.

**Supplemental Figure 2 - T cell proliferation in MIS-C is greater than in COVID-19**

(**A**) Gating scheme and representative flow cytometry plots for CD4 and CD8 T cell populations, from PBMC staining. (**B**) Frequencies of nonnaive(nn) CD4 T cell memory populations. (**C**) Frequencies of nnCD8 T cell memory populations. (**D**) Representative flow cytometry plots and quantification of Ki67+ nnCD4 T cells. (**E**) Representative flow cytometry plots and quantification of Ki67+ nnCD4 T cells. (**F**) Distribution of T cell memory subsets within HLA-DR+CD38+ nnCD4 (left) and CD8 T cells (right). (**G**) Distribution of T cell memory subsets within Ki67+ nnCD4 (left) and CD8 T cells (right). (**B-G**) Each dot represents an individual patient or subject, with adult HD in grey circles, adult COVID-19 in shades of mauve indicated by disease severity score, Ped COVID-19 in blue circles, with ARDS patients in dark blue, and MIS-C in green triangles. (**BCDE**) Significance determined by unpaired Wilcoxon test between Ped COVID-19 and MIS-C groups only, indicated by: ** $p<0.01$, or P value shown.

**Supplemental Figure 3 - NK and CD8 MAIT cell activation in MIS-C is greater than in COVID-19**

(**A**) Representative flow cytometry plots and quantification of HLA-DR and CD38 single and double-expressing NK cells from the whole blood panel. (**B**) Representative flow cytometry plots and quantification of HLA-DR and CD38 single and double-expressing CD8 MAIT from the whole blood panel. (**AB**) Each dot represents an individual patient or subject, with adult HD in grey circles, adult COVID-19 in shades of mauve indicated by disease severity score, Ped COVID-19 in blue circles, with ARDS patients in dark blue, and MIS-C in green triangles. Significance determined by unpaired Wilcoxon test between Ped COVID-19 and MIS-C groups only, indicated by: * $p<0.05$, or P value shown.

**Supplemental Figure 4 - Vascular patrolling CX3CR1+ populations in MIS-C are uniquely proliferative in MIS-C**

(**A**) Quantification of Ki67+ population within CX3CR1- and CX3CR1+ nnCD4 T cells. (**B**) Quantification of Ki67+ population within CX3CR1- and CX3CR1+ nnCD8 T cells. (**C**) Representative gating of Ki67+ population within CX3CR1- and CX3CR1+ nnCD8 T cells. (**AB**) Each dot represents an individual patient or subject, with adult HD in grey circles, adult COVID-19 in shades of mauve indicated by disease severity score, Ped COVID-19 in blue circles, with ARDS patients in dark blue, and MIS-C in green triangles. Significance determined by unpaired Wilcoxon test between Ped COVID-19 and MIS-C groups and determined by paired Wilcoxon test between CX3CR1- and CX3CR1+ within pediatric groups, indicated by: * $p <0.05$, ** $p<0.01$, *** $p<0.001$ or P value shown.

**Supplemental Figure 5 - A transition towards B cell memory subsets from naive is associated with IFNg in both MIS-C and pediatric COVID-19**

(**A**) Representative gating for naive B cells and memory subsets. (**B**) Frequencies of naive B cells. (**C**) Frequencies of memory B cell subsets from the nonnaive(nn)B cell population. (**D**) Spearman correlation for indicated cytokines with frequencies of B cell subsets. Top panel indicates correlations in the combined pediatric cohort, lower two panels represent correlations within the Ped COVID-19 and MIS-C respectively. Spearman's Rank Correlation coefficient (ρ) is indicated by square size and heat scale; significance indicated by: * $p<0.05$, ** $p<0.01$, *** $p<0.001$. (**E**) Spearman correlation for indicated cytokines with frequencies with HLA-DR+CD38+ and Ki67+ populations of nnCD4 and nnCD8 T cells, statistics and panels as in **D**. (**F**) Representative histogram of Eomes expression by PB for an adult HD (grey), Ped COVID-19 (blue) and MIS-C (green) and quantification of frequencies of Eomes+ PB. Significance determined by unpaired Wilcoxon test between pediatric cohorts indicated by: **** $p<0.001$.

**Supplemental Figure 6 - Total and activated cTfh frequencies are not associated with PB responses in either pediatric cohort.**

(**A**) Representative flow cytometry plot and quantification for cTfh. (**B**) Representative flow cytometry plot and quantification for activated cTfh. (**CDE**) Frequencies of PB versus cTfh from nnCD4 T cells (**C**), activated cTfh from nnCD4 T cells (**D**), activated cTfh from total cTfh (**E**). Non-parametric trend lines (Sen-Theil) with Spearman’s Rank Correlation coefficient (ρ) and associated P value shown. (**F**) Representative flow cytometry plot and quantification for both frequency of CXCR5+ nnCD4 T cells and CXCR5 GMFI on nnCD4 T cells. Significance determined by unpaired Wilcoxon test comparing adult HD to both pediatric cohorts and indicated by: * $p < 0.05$, ** $p<0.01$, *** $p<0.001$ or P value shown. (**A-F**) Each dot represents an individual patient or subject, with adult HD in grey circles, adult COVID in shades of mauve indicated by disease severity score, Ped COVID-19 in blue circles, with ARDS patients in dark blue, and MIS-C in green triangles. (**AB**) Significance determined by unpaired Wilcoxon test between pediatric cohorts.

**Supplemental Figure 7 - Features of CD4 T cell activation start to normalize over time, coincident with treatment and recovery from disease.**

(**A**) Selected CD4 T cell features over days since admission in Ped COVID-19 (left) and MIS-C subjects (right). Black lines connect repeat draws for individual subjects. For MIS-C, paired t-test P value is shown for the three subjects with repeat draws. Grey shading indicates the derived central 95% adult HD reference interval. (**B**) Flow cytometry plots for HLA-DR+CD38+ nnCD4 for MIS-C patients with repeat draws. (**C**) Flow cytometry plots for Ki67+ nnCD4 for MIS-C patients, as in (**B**). (**A**) Each dot represents an individual patient with Pediatric COVID-19 in blue circles (ARDS patients in dark blue) and MIS-C in green triangles.

| | Pediatric COVID-19 | | MIS-C | |
|---|---|---|---|---|
| **Demographics** | **Value (% or Range)*** | **N**** | **Value (% or Range)*** | **N**** |
| Age (Years) | 14 [0.15-18] | 16 | 9 [5.00-17] | 14 |
| Sex (F/M) | 8 (50%) / 8 (50%) | 16 | 7 (50%) / 7 (50%) | 14 |
| BMI (Age-Matched Percentile) | 65 [1-99] | 16 | 92 [5-99] | 14 |
| Race (African-American/Caucasian) | 8 (53%) / 7 (47%) | 15 | 4 (31%) / 9 (69%) | 13 |
| **Medical History** | | | | |
| Obesity | 2 (12%) | 16 | 5 (36%) | 14 |
| Diabetes Mellitus | 1 (6%) | 16 | 0 (0%) | 14 |
| Symptomatic | 13 (81%) | 16 | 14 (100%) | 14 |
| Coinfection | 5 (31%) | 16 | 1 (7%) | 14 |
| Disease Type (COVID-19 non-ARDS / COVID-19 with ARDS / MIS-C) | 12 (75%) / 4 (25%) / 0 (0%) | 16 | 0 (0%) / 0 (0%) / 14 (100%) | 14 |
| **Treatment** | | | | |
| Hydroxychloroquine | 1 (6%) | 16 | 0 (0%) | 14 |
| Remdesevir | 4 (29%) | 14 | 1 (11%) | 9 |
| Convalescent Plasma | 2 (12%) | 16 | 0 (0%) | 14 |
| Tociluzimab | 3 (19%) | 16 | 0 (0%) | 14 |
| Vasoactive Medication | 8 (50%) | 16 | 9 (64%) | 14 |
| Intravenous Immunoglobulin | 1 (6%) | 16 | 14 (100%) | 14 |
| Intravenous Immunoglobulin Before First Draw Date | 0 (0%) | 1 | 11 (79%) | 14 |
| Steroids | 6 (38%) | 16 | 13 (93%) | 14 |
| Steroids Before First Draw Date | 6 (100%) | 6 | 10 (77%) | 13 |
| **Laboratory Results** | | | | |
| Hemoglobin, 1st Research Draw | 10.3 [7.4-14.8] | 15 | 9.3 [7.6-11.8] | 14 |
| Platelet Count, 1st Research Draw | 170 [13-581] | 15 | 152 [70-537] | 14 |
| Platelet Count, Nadir | 140 [37-578] | 16 | 134 [33-194] | 14 |
| White Blood Cell Count, 1st Research Draw | 10.0 [1.1-24.2] | 15 | 11.0 [2.2-29.8] | 14 |
| Polymorphonuclear Cell Count, Absolute, 1st Research Draw | 7.8 [0.2-21.3] | 15 | 10.3 [2.0-25.8] | 14 |
| Lymphocyte Count, Absolute, 1st Research Draw | 1.8 [0.4-5.3] | 15 | 0.9 [0.1-5.7] | 14 |
| Monocyte Count, Absolute, 1st Research Draw | 0.5 [0.0-1.9] | 15 | 0.2 [0.0-1.4] | 14 |
| Eosinophil Count, Absolute, 1st Research Draw | 0.0 [0.0-0.3] | 15 | 0.0 [0.0-0.4] | 14 |
| Basophil Count, Absolute, 1st Research Draw | 0.0 [0.0-0.1] | 15 | 0.0 [0.0-0.1] | 14 |
| Lymphocyte Count, Absolute, Nadir | 0.9 [0.1-2.8] | 16 | 0.4 [0.1-1.5] | 14 |
| Sodium, Nadir | 137 [130-144] | 15 | 131 [126-140] | 14 |
| D-Dimer, Peak | 1.9 [0.2-37.4] | 10 | 4.9 [1.2-48.0] | 13 |
| Ferritin, Peak | 217 [60-16973] | 7 | 892 [253-3868] | 13 |
| Lactate Dehydrogenase, Peak | 2874 [564-15390] | 5 | 864 [457-2761] | 12 |
| Troponin, Peak | 0.1 [0.0-1.8] | 4 | 0.6 [0.0-7.4] | 13 |
| Procalcitonin, Peak | 17.62 [0.07-200.00] | 8 | 10.04 [0.24-200.00] | 11 |
| Non-Cardiac C-Reactive Protein, Peak | 21.4 [0.7-60.1] | 13 | 26.2 [6.8-40.0] | 14 |

*Values are expressed as counts (with percent of total) for discrete data or as medians [with range] for continuous data.
**N is the number of patients with non-missing annotation for each variable.

#### Supplementary Table S1: Patient Characteristics

| VENDOR | CAT# | ANTIBODY/DYE and TARGET | CLONE | DILUTION |
|---|---|---|---|---|
| BD OptiBuild | 740298 | BUV395 mouse anti-human CD45RA | HI100 | 800 |
| BD Horizon | 612943 | BUV496 mouse anti-human CD8 | RPA-T8 | 400 |
| BD Horizon | 612916 | BUV563 mouse anti-human CD19 | SJ25C1 | 200 |
| BD OptiBuild | 751572 | BUV615 mouse anti-human CD16 | 3G8 | 200 |
| BD Horizon | 612969 | BUV661 mouse anti-human CD38 | HIT2 | 400 |
| BD Horizon | 612829 | BUV737 mouse anti-human CD27 | L128 | 100 |
| BD Horizon | 612905 | BUV805 mouse anti-human CD20 | 2H7 | 800 |
| Biolegend | 329920 | BV421 mouse anti-human CD279 (PD-1) | EH12.2H7 | 100 |
| BD Horizon | 566138 | BV480 mouse anti-human IgD | IA6-2 | 50 |
| Tombo | 13-0870-T500 | Ghost Dye™ Violet 510 | NA | 400 |
| Biolegend | 300436 | BV570 mouse anti-human CD3 | UCHT1 | 25 |
| Biolegend | 356520 | BV605 mouse anti-human CD138 (Syndecan-1) | MI15 | 25 |
| Biolegend | 305642 | BV650 mouse anti-human CD95 (Fas) | DX2 | 100 |
| BD Horizon | 563163 | BV711 mouse anti-human CD21 | B-ly4 | 800 |
| BD Horizon | 566355 | BV750 mouse anti-human CD4 | SK3 | 20000 |
| BD Horizon | 564041 | BV786 mouse anti-human HLA-DR | G46-6 | 100 |
| BD Horizon | 564624 | BB515 rat anti-human CD185 (CXCR5) | RF8B2 | 50 |
| BD Horizon | 566437 | BB700 rat anti-human CD197 (CCR7) | 3D12 | 50 |
| BD Biosciences | CUSTOM | BB790 Strepavidin | NA | 400 |
| Biolegend | 341617 | Biotin rat anti-human CX3CR1 | 2A9-1 | 100 |
| Miltenyi | 130-120-716 | PE recombinant anti-human TOX | REA473 | 50 |
| Invitrogen | 61-4877-42 | PE-eFluor610 mouse anti-human EOMES | WD1928 | 100 |
| Invitrogen | GRB18 | PE-Cy5.5 mouse anti-human Granzyme B | GB11 | 2000 |
| Biolegend | 644824 | PE-Cy7 mouse anti-human T-bet | 4B10 | 800 |
| ThermoFisher | 15-9949-82 | PE-Cy5 hamster anti-human CD278 (ICOS) | C398.4A | 50 |
| CellSignalingTechnology | 6709S | AF647 rabbit anti-human TCF1/TCF7 | C63D9 | 100 |
| BD Horizon | 561277 | AF700 mouse anti-human Ki-67 | B56 | 200 |
| Biolegend | 328230 | APC/Fire750 anti-human CD39 | A1 | 50 |

**Supplementary Table 2: Panel for Peripheral Blood Mononuclear Cell Flow Cytometric Staining**

| VENDOR | CAT# | ANTIBODY/DYE and TARGET | CLONE | DILUTION |
|---|---|---|---|---|
| BD Horizon | 564071 | BUV395 mouse anti-human Ki67 | B56 | 200 |
| BD Horizon | 564804 | BUV496 mouse anti-human CD8 | RPA-T8 | 500 |
| BD Horizon | 565702 | BUV563 mouse anti-human CD45RA | HI100 | 500 |
| BD Horizon | 565069 | BUV661 mouse anti-human CD38 | HIT2 | 400 |
| BD Horizon | 564385 | BUV737 mouse anti-human CD25 | 2A3 | 200 |
| BD Horizon | 565515 | BUV805 mouse anti-human CD3 | UCHT1 | 125 |
| Biolegend | 329920 | BV421 mouse anti-human CD279 (PD-1) | EH12.2H7 | 83.3 |
| BD Horizon | 566141 | BV480 mouse anti-human CD14 | MP9 | 500 |
| Invitrogen | L34957 | LIVE/DEAD™ Fixable Aqua Dead Cell Stain Kit | NA | 15000 |
| Biolegend | 318330 | BV570 mouse anti-human CD56 | HCD56 | 200 |
| BD Horizon | 562845 | BV605 mouse anti-human HLA-DR | G46-6 | 250 |
| Biolegend | 302828 | BV650 mouse anti-human CD27 | O323 | 200 |
| Biolegend | 302044 | BV711 mouse anti-human CD16 | 3G8 | 666.7 |
| BD OptiBuild | 747111 | BV750 rat anti-human CD185 (CXCR5) | RF8B2 | 500 |
| Biolegend | 302240 | BV785 mouse anti-human CD19 | HIB19 | 125 |
| BD Horizon | 555401 | FITC mouse anti-human CD15 | HI98 | 50 |
| BD OptiBuild | 746472 | BB700 mouse anti-human CD103 | Ber-ACT8 | 500 |
| BD Biosciences | CUSTOM | BB790 mouse anti-human CD4 | SK3 | 1250 |
| BD OptiBuild | 746472 | PE mouse anti-human CD123 | 9F5 | 400 |
| BD Horizon | 562397 | PE-CF594 mouse anti-human CD127 | HIL-7R-M21 | 200 |
| Biolegend | 310908 | PE-Cy5 mouse anti-human CD69 | FN50 | 500 |
| eBioscience | 35-0114-82 | PE-Cy5.5 mouse anti-human CD11c | N418 | 100 |
| Biolegend | 354912 | PE-Cy7 mouse anti-human CD21 | BU32 | 2000 |
| BD Horizon | 550968 | APC mouse anti-human CD161 | DX12 | 10 |
| BD Horizon | 560566 | AF700 mouse anti-human CD45 | HI30 | 83.3 |
| Biolegend | 353212 | APC-Cy7 mouse anti-human CD197 (CCR7) | G043H7 | 66.7 |

#### Supplementary Table 3: Lineage Panel for Whole Blood Flow Cytometric Staining

#### Supplemental Figure 1

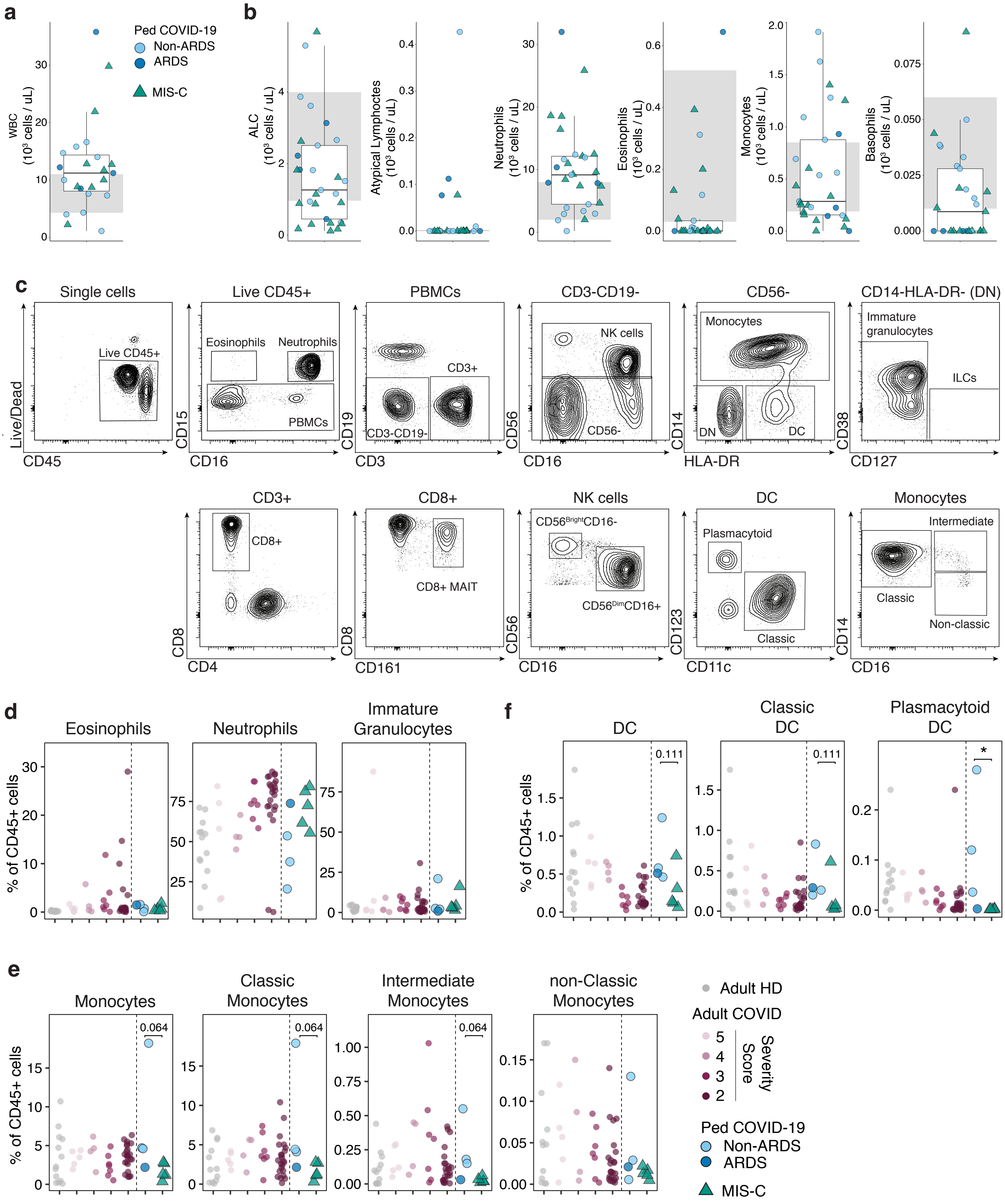

### Supplemental Figure 2

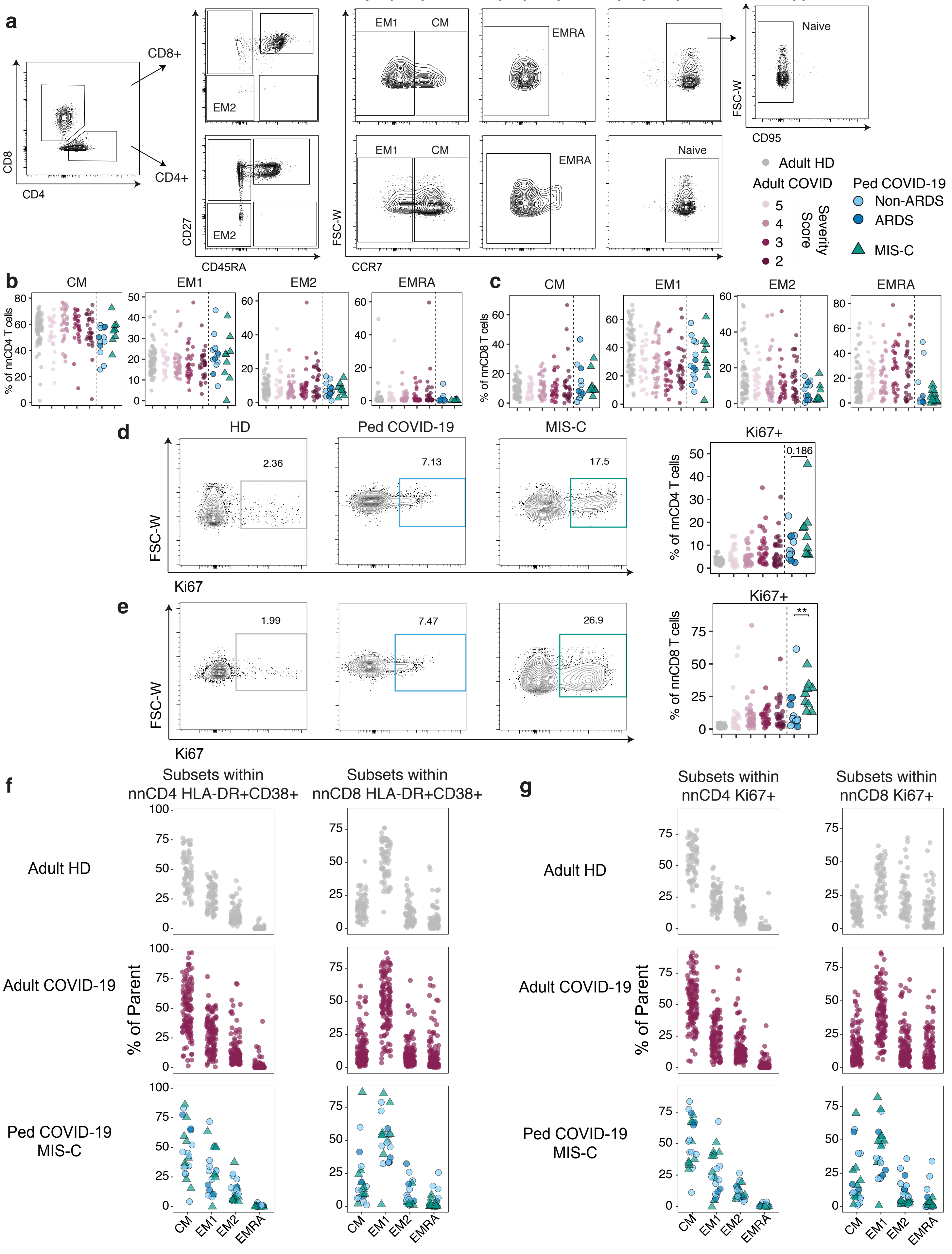

### Supplemental Figure 3

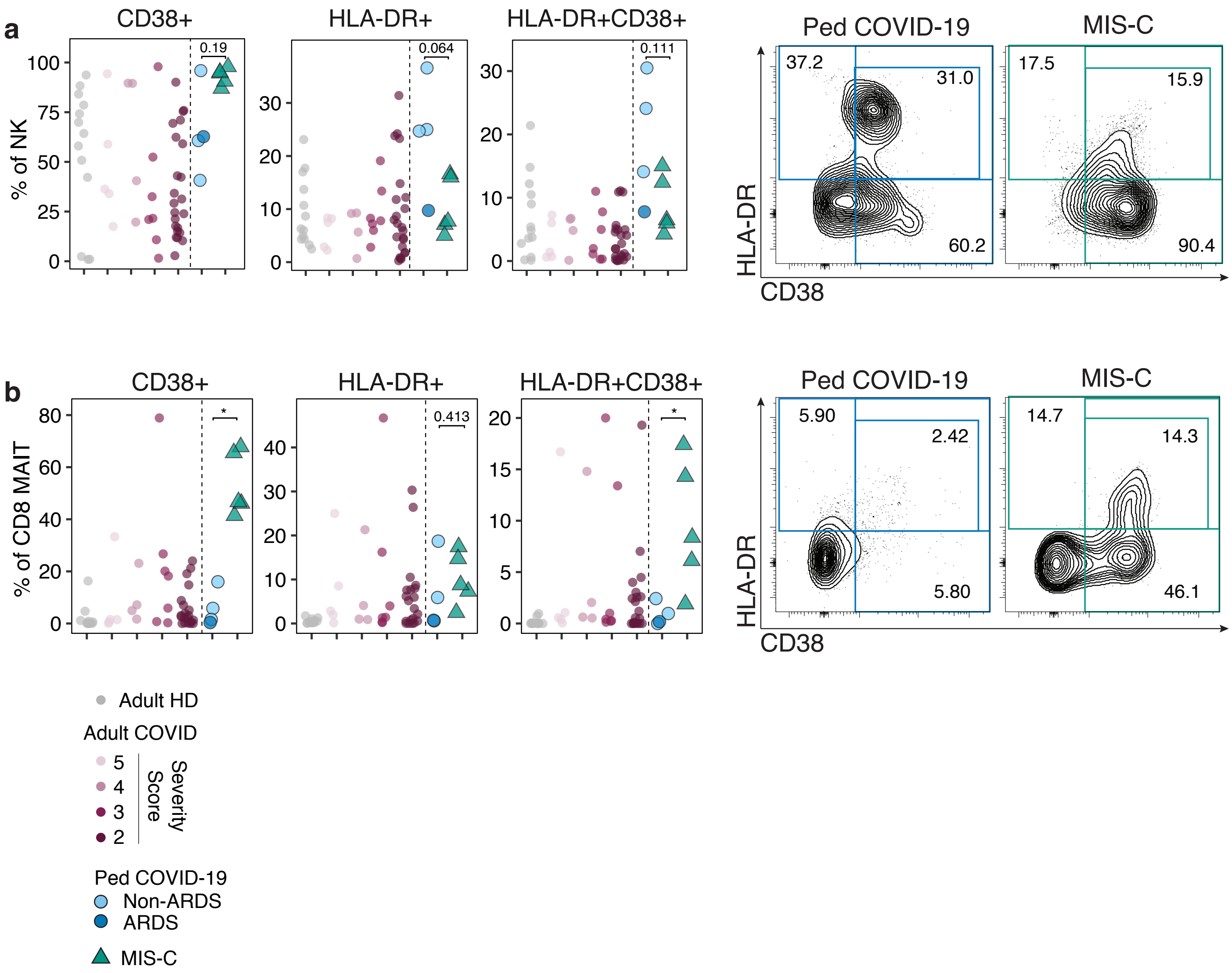

#### Supplemental Figure 4

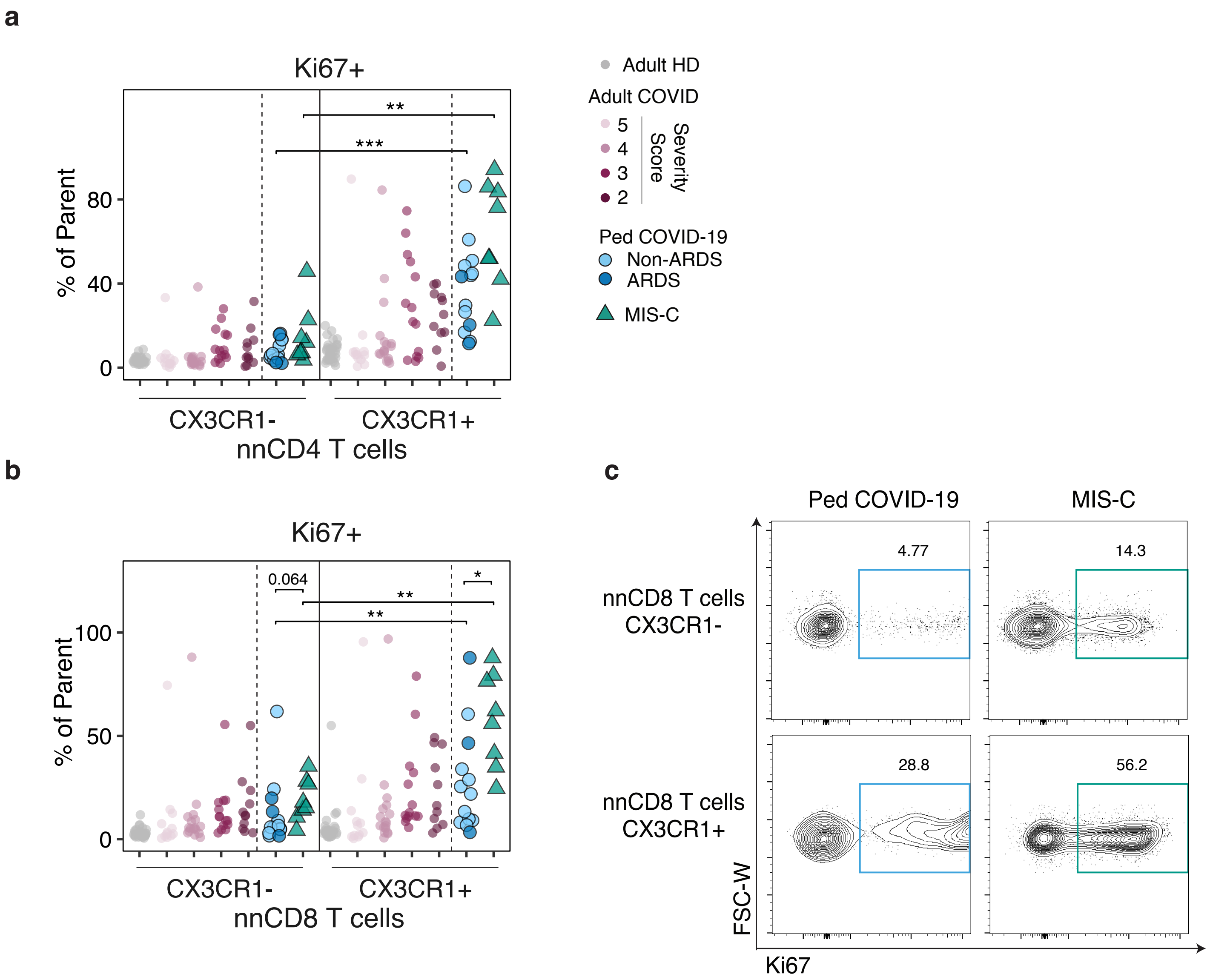

**Supplemental Figure 5**

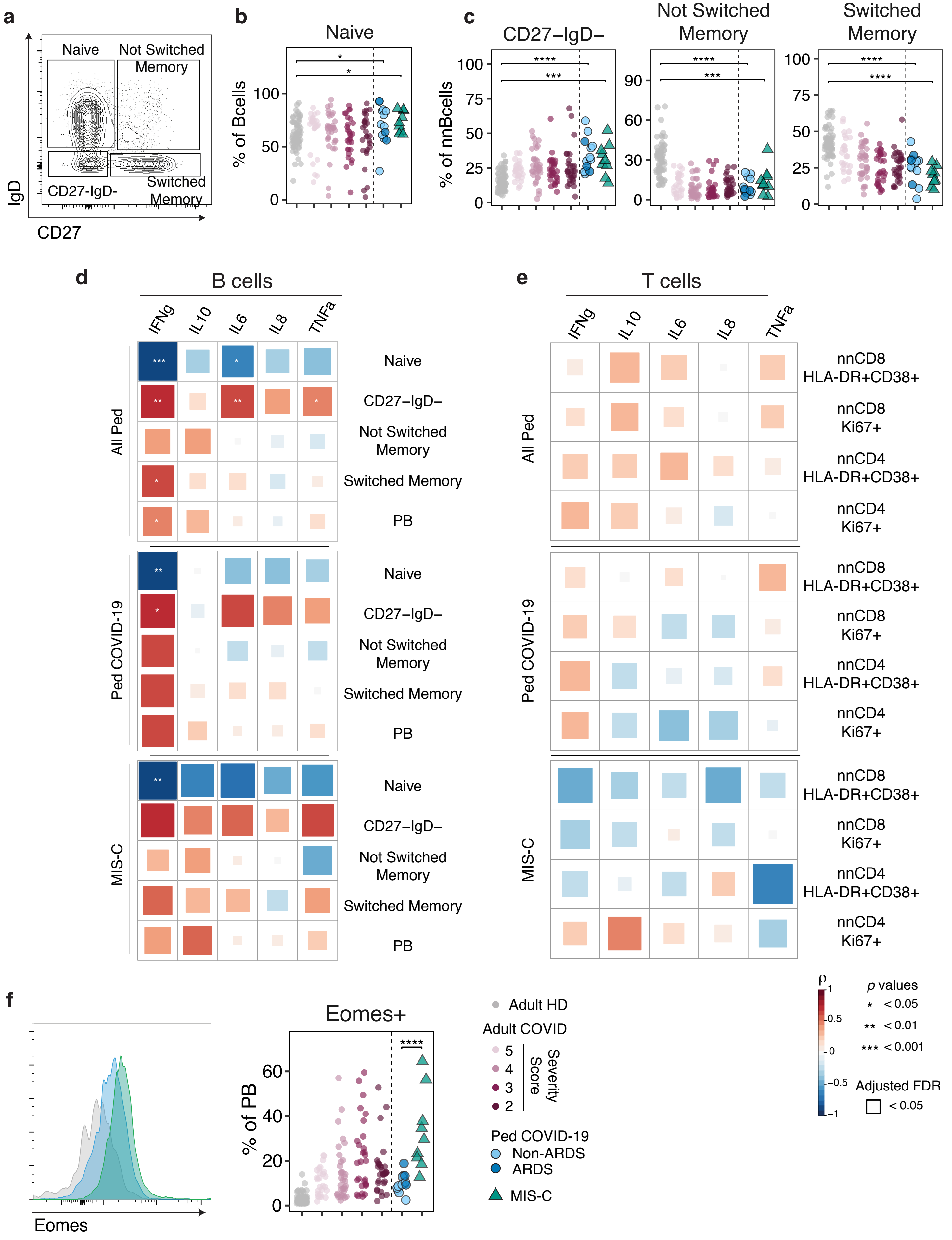

#### Supplemental Figure 6

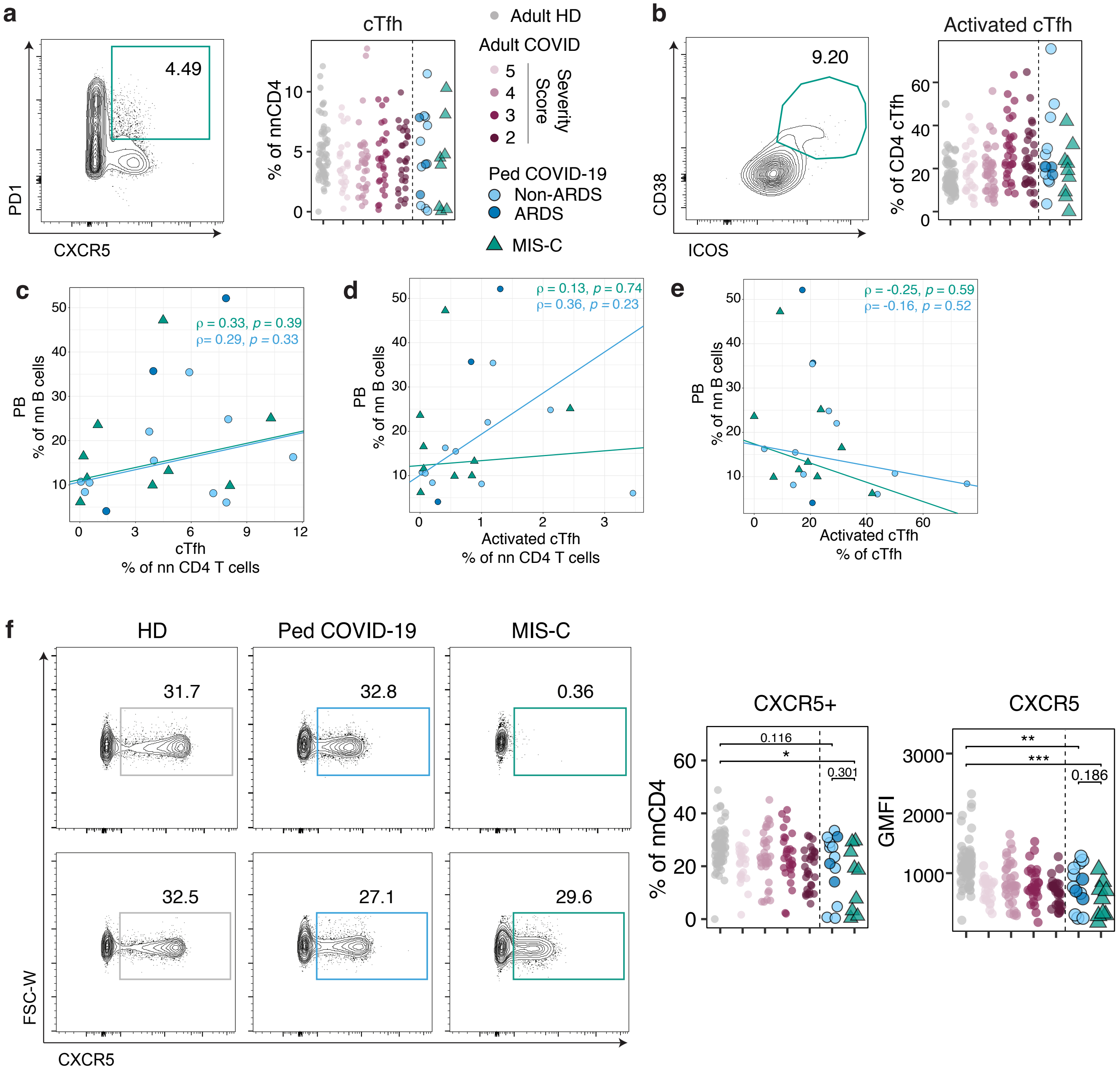

#### Supplemental Figure 7

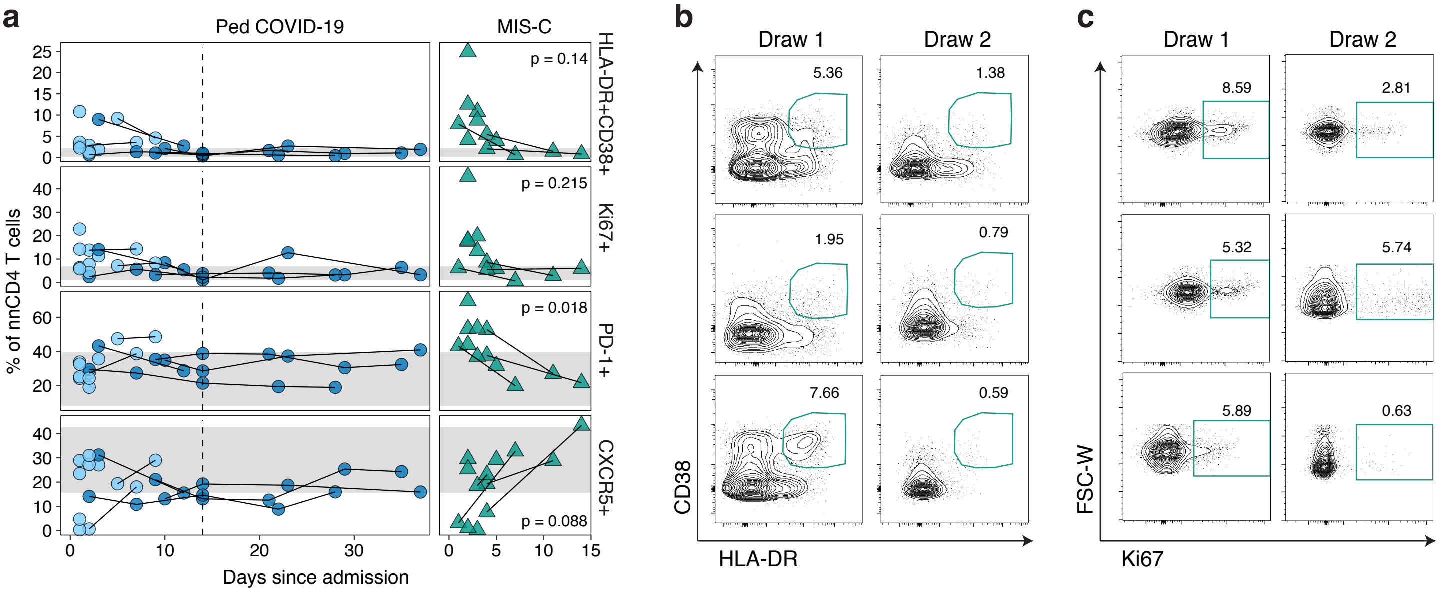
